# IDH1-dependent m6A methylation defines transcriptomic heterogeneity in glioma

**DOI:** 10.1101/2024.09.24.24314089

**Authors:** Syeda Maheen Batool, Hanna Lee, Koushik Muralidharan, Saad Murtaza Khan, Ana K. Escobedo, Denalda Gashi, Kesli Faber, Emil Ekanayake, Tiffaney Hsia, Yana Al-Inaya, Aishwarya Kosgi, Julie J. Miller, Daniel P. Cahill, Gavin P. Dunn, Bryan D. Choi, Allegra S. Petti, Bob S. Carter, Leonora Balaj

## Abstract

Gliomas are biologically heterogeneous brain tumors with marked differences in clinical behavior based on the IDH1 mutation status. While epigenetic dysregulation is well characterized, the contribution of RNA modifications, particularly N6-methyladenosine (m6A), remains underexplored. Using direct RNA nanopore sequencing of patient-derived gliomas, we generated the first isoform-resolved m6A maps across IDH1-mutant and wild-type tumors. IDH1-mutant gliomas exhibited globally elevated m6A methylation, along with increased expression of methyltransferases (METTL3, METTL14) and stabilizing readers (YTHDF3). In contrast, wild-type glioblastomas showed enhanced expression of m6A erasers (ALKBH5, FTO) and RNA decay factors (YTHDF2). These subtype-specific differences in m6A architecture impacted transcript stability, isoform usage, and gene expression. Isoform-level analyses revealed stronger prognostic associations than gene-level parameter, including for IGF2BP2-202, PUF60-202, and GLUL-203. Our study establishes m6A as a critical, subtype-specific layer of RNA regulation in glioma with clinical and therapeutic implications.

## Introduction

Gliomas are the most common primary brain tumors in adults^1^. Despite significant progress in understanding their molecular landscape, the prognosis for patients with high-grade gliomas, particularly glioblastoma (GBM), remains dismal, with median survival under two years^1,2^. In contrast, lower-grade gliomas carrying mutations in isocitrate dehydrogenase 1 (IDH1), including astrocytomas and oligodendrogliomas, demonstrate slower progression and significantly improved outcomes^3^. These clinical differences are reflected in distinct molecular phenotypes and have established IDH1 mutation status as a critical determinant in glioma classification and management^3,4^.

A hallmark of IDH1-mutant gliomas is widespread epigenetic reprogramming, driven by accumulation of the oncometabolite D-2-hydroxyglutarate (2-HG), which inhibits α-ketoglutarate (α-KG)–dependent enzymes including DNA and histone demethylases^5^. This leads to global DNA hypermethylation and altered chromatin states that contribute to transcriptional silencing and tumor evolution^5^. While these DNA-level changes are well described, much less is known about how post-transcriptional mechanisms, specifically RNA modifications, shape glioma biology. Among the >170 known RNA modifications, N6-methyladenosine (m6A) is the most abundant internal mark on mRNA and plays essential roles in RNA splicing, translation, stability, and degradation^5–7^. The distinct groups of binding proteins, widely known as “m6A regulators” (readers, writers, and erasers) cooperatively interact to deposit, remove, and recognize m6A. The dynamic and reversible changes in m6A influence key stages of RNA life cycles including splicing, nuclear export, degradation, and translation (ref). Importantly, m6A marks are deposited and removed by enzymes that rely on α-KG, such as the “writers” METTL3/METTL14 and the “erasers” FTO and ALKBH5, suggesting that IDH1 mutation may also remodel the glioma epitranscriptome^8^.

Emerging evidence links m6A to diverse cancer phenotypes including proliferation, therapy resistance, and immune evasion, yet its role in glioma remains incompletely understood. Among the m6A regulators, studies have described METTL3 dependent regulation of RNA processing and stabilization in glioma stem cells (GSCs) with downstream impact on gliomagenesis ^9,10^. Prior studies have mostly relied on methylated RNA immunoprecipitation sequencing (MeRIP-seq), which lacks isoform resolution and site specificity^11^. In gliomas, where alternative splicing and noncoding isoform expression are pervasive, this represents a critical limitation^11,12^. Moreover, the interplay between m6A methylation, transcript structure, immune signaling, and clinical outcomes across glioma subtypes has not been systematically examined in patient-derived tissue. Long-read RNA sequencing technologies, such as those from Oxford Nanopore Technologies, overcome these limitations by providing single-nucleotide resolution of m6A modifications^13^. Additionally, recent advancements in deep learning-based models m6A prediction models allow for high resolution and transcriptome-wide coverage of m6A sites^14^.

In this study, we profile the m6A RNA landscape in IDH1 mutant gliomas (n=8) and IDH1-wild-type glioblastomas (GBM, n=6). Our approach utilized the direct RNA sequencing platform of Nanopore, enabling full-length, single-molecule transcript analysis while preserving native RNA modifications. We applied the neural network model m6Anet to predict m6A modifications at single-nucleotide resolution at the site, transcript, and gene levels. By integrating m6A mapping with transcript region annotation, gene expression, isoform usage, and RNA biotype information, we characterize the post-transcriptional regulatory roles of m6A across glioma subtypes. Finally, we evaluate the diagnostic and prognostic potential of m6A modifications to assess their clinical relevance in glioma. Together, our study positions m6A as a critical, subtype-specific layer of gene regulation in glioma, with implications for RNA biology, biomarker discovery, and therapeutic development.

## Results

### IDH1 mutation defines a distinct m6a epitranscriptomic landscape in glioma

We performed transcriptome-wide m6A mapping in *n=14* glioma tumor RNA samples using Nanopore direct RNA sequencing (**Supplementary Figure 1a-b**; **see Methods).** Study population included IDH1 mutant gliomas (n=8, Astrocytoma [AA]), n=6; Oligodendroglioma [OO], n=2) and IDH1 wild-type glioblastoma (GBM, n=6). In addition, n = 2 entry cortex brain tissue samples were included (**see Methods**). m6A RNA modifications were predicted using m6Anet^14^(**see Methods, Supplementary Figure 1c**).

Overall, a higher burden of m6A sites, transcripts, and genes was observed in the IDH1 mutant gliomas (OO, AA) compared to the IDH1 wild-type GBM group (**Fig. 1a-c, left panel**). Among all subtypes, OO had the highest percentage of m6A modified sites (3.0%, n = 8,139), transcripts (22.9%, n = 4,611), and genes (26.5%, n = 1,614) (**Fig. 1b, left panel**). This was followed by AA (sites: 1.9%, n = 850; isoforms: 14.1%, n = 536; genes: 16.2%, n = 181) and GBM (sites: 1.6%, n = 421; isoforms: 11.4%, n = 290; genes: 12.9%, n =86) (**Fig. 1a-c, left panel**) (**Supplementary Figure 5a**).

**Figure 1.**
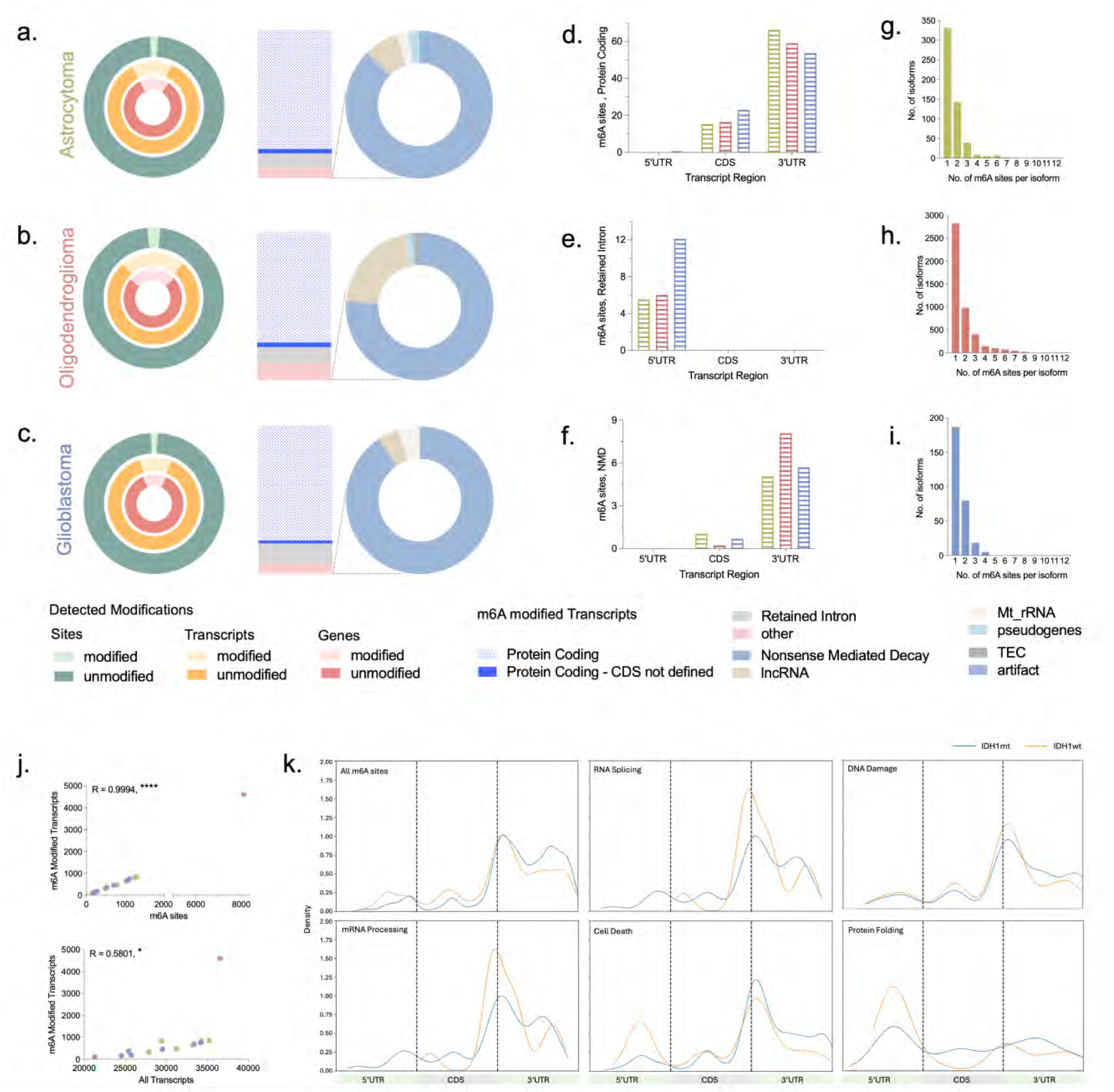
m6A modified sites, transcripts, and genes in IDH1 mutant and wild-type gliomas. **(a-c)** Pie charts (left) showing the prevalence of m6A modified sites (green), isoforms (orange), and genes (red) in each glioma subtype (AA, Astrocytoma (green); OO, Oligodendroglioma (red); GBM, Glioblastoma (blue)). Stacked bars and accompanying pie charts (right) depict the biotype distribution of m6A-modified isoforms, including protein-coding and less abundant RNA biotypes. (d-f) Proportion of m6A sites mapped to 5’UTR, CDS and 3’UTR regions within (d) protein-coding, (e) retained intron, and (f) nonsense-mediated decay (NMD) isoforms across glioma subtypes (AA: green, OO: red, GBM: blue). **(g-i)** Distribution of single-site or multi-site (>1) m6A-modified isoforms in (g) AA, (h) OO, and (i) GBM. **(j)** Correlation plot showing: (top) number of m6A-modified isoforms vs. number of sites per patient (probability >0.9); and (bottom) total isoforms detected vs number of m6A-modified isoforms. **(k)** Metagene plots showing m6A site distribution across isoform regions in IDH1 mutant (IDH1mt) and wild-type (IDHwt) glioma across pathways including RNA splicing, DNA Damage, mRNA Processing, Cell Death, and Protein Folding.

The m6A RNA biotypes were differentially distributed across the glioma subtypes, (**Fig. 1a-c, right panel**) despite an overall similar RNA biotype distribution (**Supplementary Figure 4a**). Overall, across the glioma subtypes, protein coding RNA was the most frequently m6A modified RNA biotype. However, AA had the highest prevalence of m6A modified protein coding RNA (80.2%) when compared to OO (74.7%) and GBM (77.6%) (**Fig. 1a-c, right panel**). In contrast, retained intron (RI) RNA was more frequently modified in the GBM (13.1%) when compared to IDH1 mutant glioma subtypes (AA: 9.1%, OO: 10.7%). Among non-protein coding RNA biotypes, OO group had highest enrichment of m6A modified nonsense-mediated decay (NMD) RNA (8.7%, AA: 6.5%, GBM: 6.6%) and lncRNA (2.3%, AA: 0.6%, GBM 0.0%) (**Fig. 1a-c, right panel**).

We then proceeded to further investigate the glioma subtype specific differences in the prevalence of m6A modified RNA biotypes (protein coding, retained intron, NMD) at the transcript region level (5’UTR, CDS, 3’UTR, **Fig. 1d-f**). Protein coding RNA (**Fig. 1d**) overall was most frequently targeted for m6A modification in the CDS and 3’UTR regions. Interestingly, IDH1 mutant gliomas had a distinct pattern with higher m6A sites in the 3’UTR (AA: 66.2%, OO: 59.1%) when compared to the GBM subtype (53.7%). In contrast, GBM subtype had a higher number of m6A sites localized to the CDS region (23.0%) when compared to AA (15.3%) and OO (16.5%) (**Fig. 1d**). Retained Intron m6A RNA was exclusively modified in the 5’UTR region with overall 2-fold higher m6A sites noted in the GBM category (12.1%, AA: 5.5%, OO: 6.0%) (**Fig. 1e**). Finally, NMD RNA was seen to be m6A modified preferentially in the 3’UTR region followed by CDS region across all the glioma subtypes (**Fig. 1f**). However, OO was seen to have a slightly higher prevalence of NMD m6A sites in the 3’UTR region (8.1%, AA: 5.1%, GBM: 5.7%) and lower enrichment in the CDS region (0.23%) when compared to AA (1.1%) and GBM (0.7%) (**Fig. 1f**). The length distribution of all reads and full length reads highlighted a higher abundance of shorter RNA fragment (0-500bp) in the GBM subtype (32.6%, AA: 36.9%, OO: 38.2%, GBM) and a higher abundance of longer RNA fragments (1001-5000bp) in the AA subtype (0.036%, AA: 0.033%, OO: 0.019%, GBM; **Supplementary Figure 4b**). The lengths of the poly A tails were similar across groups (**Supplementary Figure 4c**). Fusions were also detected albeit only two passed the high confidence filters (**Supplementary Figure 4d**).

Next, we quantified the prevalence of single- vs. multi- site m6A methylated RNA isoforms in AA (**Fig. 1g**), OO (**Fig. 1h**), and GBM (**Fig. 1i**). Consistent with previous findings of overall higher m6A RNA levels in IDH1 mutant gliomas, we observed a remarkably higher number of single and multi-methylated m6A sites in AA and OO compared to GBM (**Fig. 1g-i**). Not only GBM isoforms were predominantly single-site methylated, the overall prevalence of single-site isoforms was much lower in GBM (n = 149) compared to AA (n = 331) and OO (n = 2826) (**Fig. 1g-i**). Comparison of the multi-site methylated isoforms demonstrated important differences. In AA group, maximum 7 m6A sites per isoform (**Fig. 1g**) were seen compared to only 3 m6A sites per isoform in GBM (**Fig. 1i**). Consistent with previous findings of m6A hypermethylation, OO had the highest prevalence of single- and multi- methylated isoforms (up to 12 sites per isoform) (**Fig. 1h**).

In our analysis of glioma tumor tissue, we identified a significant number of m6A motifs (probability score ≥ 0.9), categorized into canonical DRACH motifs (GGACT, GAACA, GAACT) and non DRACH motifs (GCACA, GGACC, AGACT) (**Supplementary Figure 5i**). No remarkable differences were observed in the nucleotide composition and probability distribution of the DRACH motifs across the glioma subtypes (**Supplementary Figure 5i**). Quantification of the non-DRACH kmers demonstrated a higher enrichment of GGACC in IDH mutant gliomas (AA: 8.6%, OO: 10.7%) vs. GBM (3.3%) (**Supplementary Figure 5i**). In addition to differences in prevalence, glioma subtypes also demonstrated distinct transcript region specific localization patterns of the identified motifs (**Supplementary Figure 5e**). GGACA and GAACT motifs were more likely to be enriched in the 3’UTR region of the IDH1 mutant group (**Supplementary Figure 5e**). The GBM group demonstrated a higher 5’UTR enrichment of the GAACT motif (**Supplementary Figure 5e**).

Our transcript region (5’UTR, CDS, 3’UTR) analysis was expanded to integrate the chromosomal location of common m6A targets across the glioma subtypes. Overall, we did notice that 5’UTR specific m6A sites most frequently localized to the chromosomes 6 and 9 in the GBM group (**Supplementary Figure 5b-d, 5f**). In contrast, CDS and 3’UTR m6A sites were more widely distributed across the glioma subtypes **(Supplementary Figure 5f)**. Within the 5’UTR region, we stratified the m6A sites by the RNA biotype and noted that the highest difference was seen in the retained intron RNA (**Supplementary Figure 5h**), consistent with earlier findings. To identify potential differences in the m6A targets at the chromosomal level, we plotted the chromosomal targets of all the glioma subtypes where the prevalence of m6A sites differs by ≥ 0.5% between any two groups. The resulting data points highlight the chromosomes where glioma subtype specific m6A prevalence differs from 0.5% to 5.0% (**Supplementary Figure 5g**). The resulting analysis demonstrated a distinct pattern of m6A localization on the chromosome unique to the IDH glioma subtype. The GBM group was seen to have a symmetrical distribution along the shorter and longer chromosomes. However, both the IDH mutant glioma subtypes (AA, OO) had a higher m6A enrichment in the shorter chromosomes (**Supplementary Figure 5g**).

Correlation analysis revealed a strong positive relationship between m6A site burden and the number of modified isoforms per patient (**Fig. 1j, top panel**). Likewise, a higher total number of detected isoforms was associated with greater m6A modification (**Fig. 1j, bottom panel**), supporting the presence of coordinated epitranscriptomic regulation.

Finally, we assessed m6A distributions in functional ontologies (**Fig. 1k**). In IDH1 wild type gliomas, m6A peaks localized around the stop codon in pathways related to RNA splicing and mRNA processing (**Fig. 1k**). In contrast, cell death and protein folding transcripts were preferentially modified at the 5’UTR (**Fig. 1k**). This subtype-specific m6A landscape suggests functional specialisation in RNA fate and regulation.

### m6A methylation differentially shapes transcript localization and gene expression across glioma subtypes

To correlate individual gene expression with m6A RNA modifications, we focused our analysis on differential methylation quantification of 226 transcripts (67 genes) commonly modified across all three glioma subtypes: AA, OO and GBM (**Fig. 2a**). We then quantified m6A methylation per transcript by first summing the m6A modification ratios of all sites detected per isoform and subsequently normalizing by the transcript length to calculate a weighted mod ratio (**see Methods, Supplementary Figure 6a**). This enabled identification of hypermethylated (Log_2_FC > 0) or hypomethylated (Log_2_FC < 0), which we subsequently correlated with gene expression levels (**Fig. 2c-e**). Three pairwise comparisons were made: AA vs GBM, OO vs GBM, AA vs OO (**Fig. 2c-e**). Across the 226 commonly modified transcripts, a significantly higher number of multi-site methylated transcripts were seen in IDH1 mutant gliomas (AA, OO) compared to GBM (**Supplementary Figure 6b**). We also observed a slight positive correlation between transcript length, number of m6A sites, and modification ratio up to 2500 bp (**Supplementary Figure 6c-d**).

**Figure 2.**
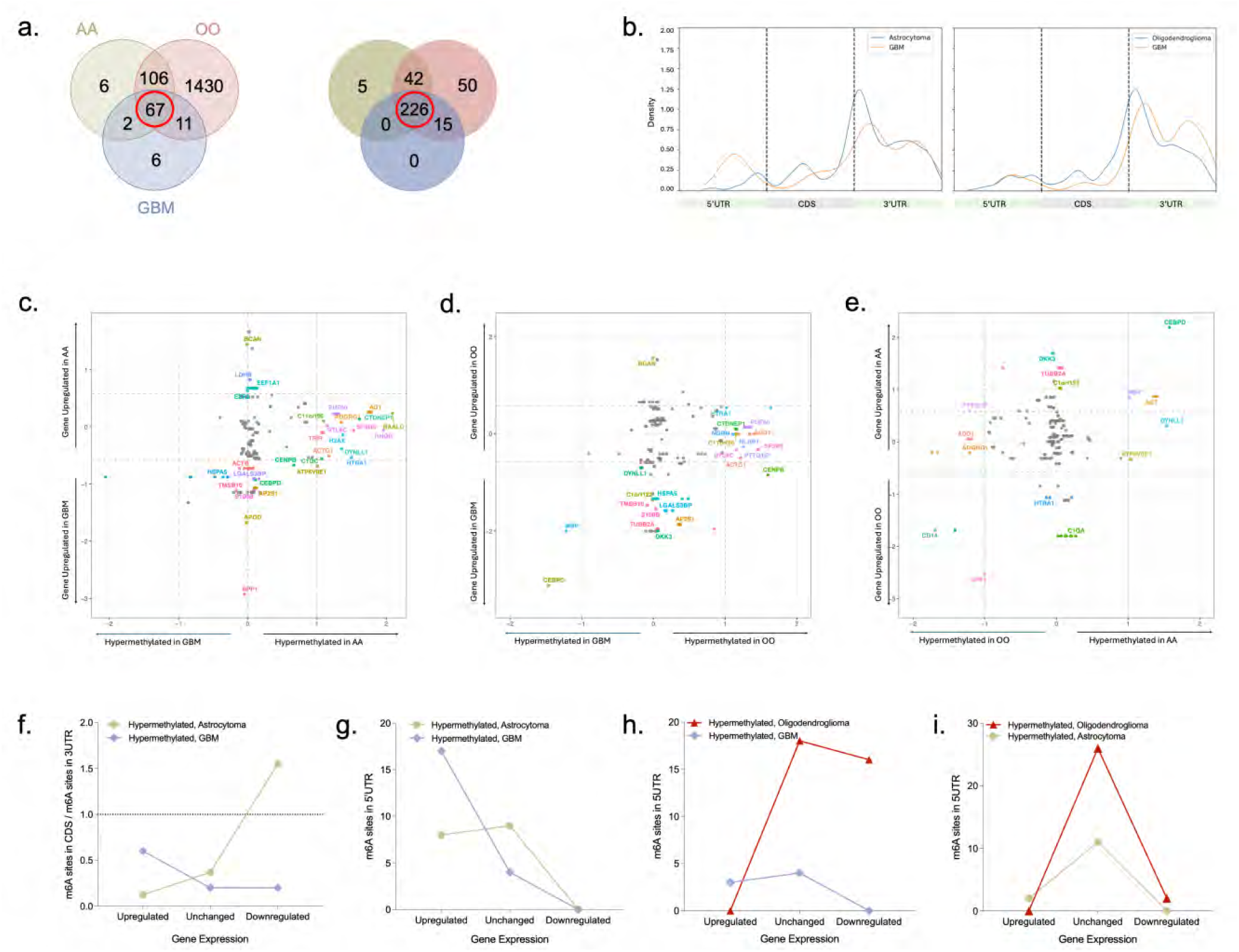
Transcript-level analysis of hyper- and hypomethylated m6A isoforms. **(a)** Venn Diagrams showing the overlap of m6A modified genes (left) and isoforms (right) across glioma subtypes (AA, OO, GBM). A total of 226 commonly modified isoforms were used for downstream analysis. **(b)** Metagene plots showing isoform-region distribution of m6A sites in hypermethylated isoforms for each subtype. **(c-e)** Scatter plots integrating m6A methylation (x-axis, log2 fold change of weighted mod ratio) and gene expression (y-axis, log2 fold change) for common m6A-modified isoforms in each comparison: (c) AA.vs GBM, (d) OO vs GBM, and (e) AA vs OO. Dashed lines indicate threshold for significance. **(f)** Ratio of m6A site density in the CDS vs 3’UTR for hypermethylated protein-coding transcripts stratified by gene expression direction (up, down, unchanged). **(g-i)** Number of m6A sites in the 5’UTR region of hypermethylated retained intron isoforms, stratified by direction of gene expression, for each comparison.

Transcriptomic localization of m6A sites in hypermethylated transcripts revealed glioma subtype specific patterns (**Fig. 2b**). In GBM, m6A sites were enriched in the 5’UTR region, while in AA there was predominant enrichment in the CDS region and near the stop codon (**Fig. 2b, left panel**). A similar comparison between OO and GBM showed enrichment near the stop codon in OO and a shift toward the tail end of the 3’UTR region in GBM (**Fig. 2b, right panel**).

Importantly, changes in methylation did not always correspond to changes in gene expression. For example, in AA vs. GBM comparison, 49 genes exhibited hypermethylated transcripts in AA without significant differences in gene expression (**Fig. 2c**). Similar trends were observed in OO vs. GBM (53 genes, **Fig. 2d**) and AA vs. OO (59 genes, **Fig. 2e**), suggesting post-transcriptional regulation that occurs independently of gene-level expression changes.

Despite this, m6A hypermethylation was associated with both upregulation and downregulation of gene expression. At the transcript level, these m6A methylation effects were observed across both protein coding retained intron RNA biotypes, sometimes within isoforms of the same gene (**Supplementary Table 1, Supplementary Figure 7b-d**).

To further investigate the mechanisms underlying the regulatory role of m6A, we analyzed region specific localization of m6A sites across hypermethylated transcripts, stratified by biotypes and direction of gene expression (upregulated, unchanged, and downregulated) (**Fig. 2f-i, Supplementary Figure 7a**). Our findings demonstrate that m6A mediated regulation of gene expression was subtype specific. In AA, hypermethylation of protein-coding transcripts was more frequently associated with gene upregulation compared to GBM (**Fig. 2f**). Conversely, in GBM, hypermethylation of RI transcripts corresponded to high gene expression (**Fig. 2g**). Furthermore, the relative distribution of m6A sites in the CDS and 3’UTR regions was predictive of gene expression. In AA, higher 3’UTR/CDS methylation was associated with increased gene expression compared to GBM (**Fig. 2f**). In contrast, in GBM, m6A enrichment in CDS/3’UTR correlated with gene upregulation **(Fig. 2f)**. Notably, the number of sites in the 5’UTR region of RI transcripts was higher in GBM, correlating with elevated gene expression when compared to AA and OO (**Fig. 2g- h**). In contrast, minimal regional differences were detected between AA and OO, both IDH1 mutant subtypes. Specifically, m6A enrichment in RI transcripts did not contribute to a significant change in gene expression in either direction (**Fig. 2i**).

### m6a-modified upregulated genes reveal divergent functional programs in IDH1 mutant and wild-type gliomas

We next performed a differential gene expression analysis (**Fig. 3a, see Methods**) across three comparison groups: (i) AA vs. GBM, (ii) OO vs. GBM, and (iii) AA vs. OO. Overall, n = 212 genes were found to be significantly upregulated in IDH1 mutant gliomas (AA vs. GBM, OO vs. GBM) and n = 387 genes upregulated in GBM (**Fig. 3a**). To identify functional consequences of m6A modification on gene expression, we filtered the upregulated genes in each group to include those with predicted m6A modification in at least one associated isoform (**Fig. 3b**). Of the 212 genes upregulated in the IDH1 mutant group, n = 68 genes were also m6A modified (**Fig. 3b**). In contrast, of the 387 genes upregulated in IDH1 wild-type glioma, only n = 17 genes were m6A modified (**Fig. 3b**).

**Figure 3.**
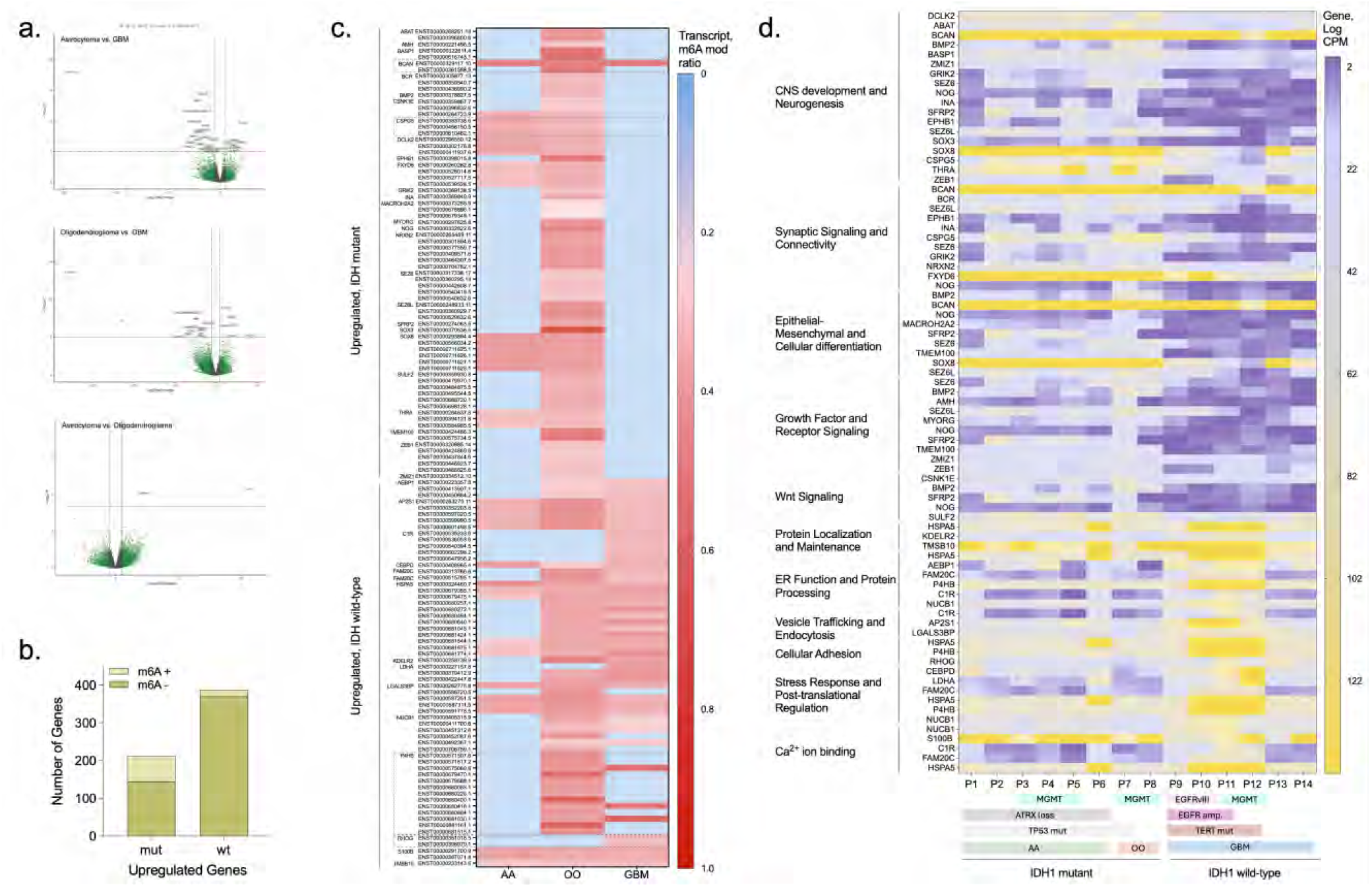
Functional functional enrichment of m6a-modified upregulated genes in IDH1 mutant and wild-type gliomas. **(a)** Volcano plots showing differentially expressed genes (DEGs) from each pairwise comparison: AA vs. GBM (top), OO vs. GBM (middle), and AA vs. OO (bottom). Significant DEGs (Log2 FC > 0.58 or < -0.58) are highlighted in red. **(b)** Stacked plot quantifying the number of upregulated genes in IDH1 mutant (mut) and IDH1 wild-type (wt) gliomas that are also m6A modified (light green). **(c)** Heatmap displaying the weighted m6A modification ratio per isoform in upregulated, m6A-modified genes from the mutant (n=68) and wild-type (n=17) group. **(d)** Heatmap of top enriched gene ontology (GO) terms among m6A modified upregulated genes. The y-axis denotes the enriched functional pathway, and the associated genes identified in each ontology term. Pathway enrichment is plotted by patient (rows) and associated gene expression (counts per million, columns), stratified by IDH1 status and key molecular features (EGFR, TERT, ATRX, TP53, MGMT).

We next quantified the m6A weighted modification ratio per isoform (**see Methods**; **Fig. 3c**). Importantly, of the 17 upregulated and m6A modified genes in the IDH1 wild-type group, only 2 genes (C1R, RHOG) were shown to be uniquely modified in GBM (**Fig. 3c**). However, the majority of the upregulated genes in the IDH1 mutant group were uniquely modified in IDH mutant gliomas, particularly OO. Finally, of the upregulated genes in GBM that were commonly modified in IDH mutant and wild-type gliomas, both hyper- and hypo- methylated isoforms were identified in GBM relative to AA and OO (**Fig. 3c**).

The m6A modified upregulated genes in IDH1 mutant and wild type groups were then further analysed to identify key downstream target pathways. The results from ontology analysis (**see Methods**) demonstrated signaling and regulatory pathways unique to each group (**Fig. 3d**). Overall, pathways enriched in IDH1 mutant gliomas included CNS development and neurogenesis, synaptic signaling, cell differentiation, growth factor and Wnt signaling. In contrast, pathways enriched in IDH1 wild-type gliomas included protein localization and maintenance, endoplasmic reticulum function and protein processing, vesicle trafficking, cellular adhesion, stress response, and Ca^2+^ ion binding (**Fig. 3d**).

### Differential isoform usage and m6A regulation drive functional transcriptomic divergence in astrocytoma and glioblastoma

Differential isoform usage across biotypes can play a critical role in regulating gene expression. We study this in our glioma cohort using a multifaceted approach combining differential gene expression (DEG), differential isoform expression (DEI), and differential isoform usage (DIU) using DeSeq and *IsoformSwitchAnalyzerR* (**see Methods**).

Clonal subtype analysis identified a total of 159 statistically significant isoform switches across 139 genes: n = 148 between AA and GBM, n = 11 between AA and OO (**Fig. 4a, Supplementary Figure 8a**). Among the 148 isoforms with significant differential usage (dIF) between AA and GBM, high usage isoforms were similarly distributed between AA (n = 77) and GBM (n = 71) (**Fig. 4a**). However, the underlying biotype distribution differed remarkably (**Fig. 4b**). AA had a greater number of high usage protein coding isoforms (n = 67) compared to GBM (n = 41). In contrast, retained intron (RI) isoforms exhibited higher usage in GBM (n = 22 vs n = 3 in AA). Finally, a similar distribution of high usage lncRNA isoforms was identified in both subtypes (AA: n = 6, GBM: n = 3, **Fig. 4b**).

**Figure 4.**
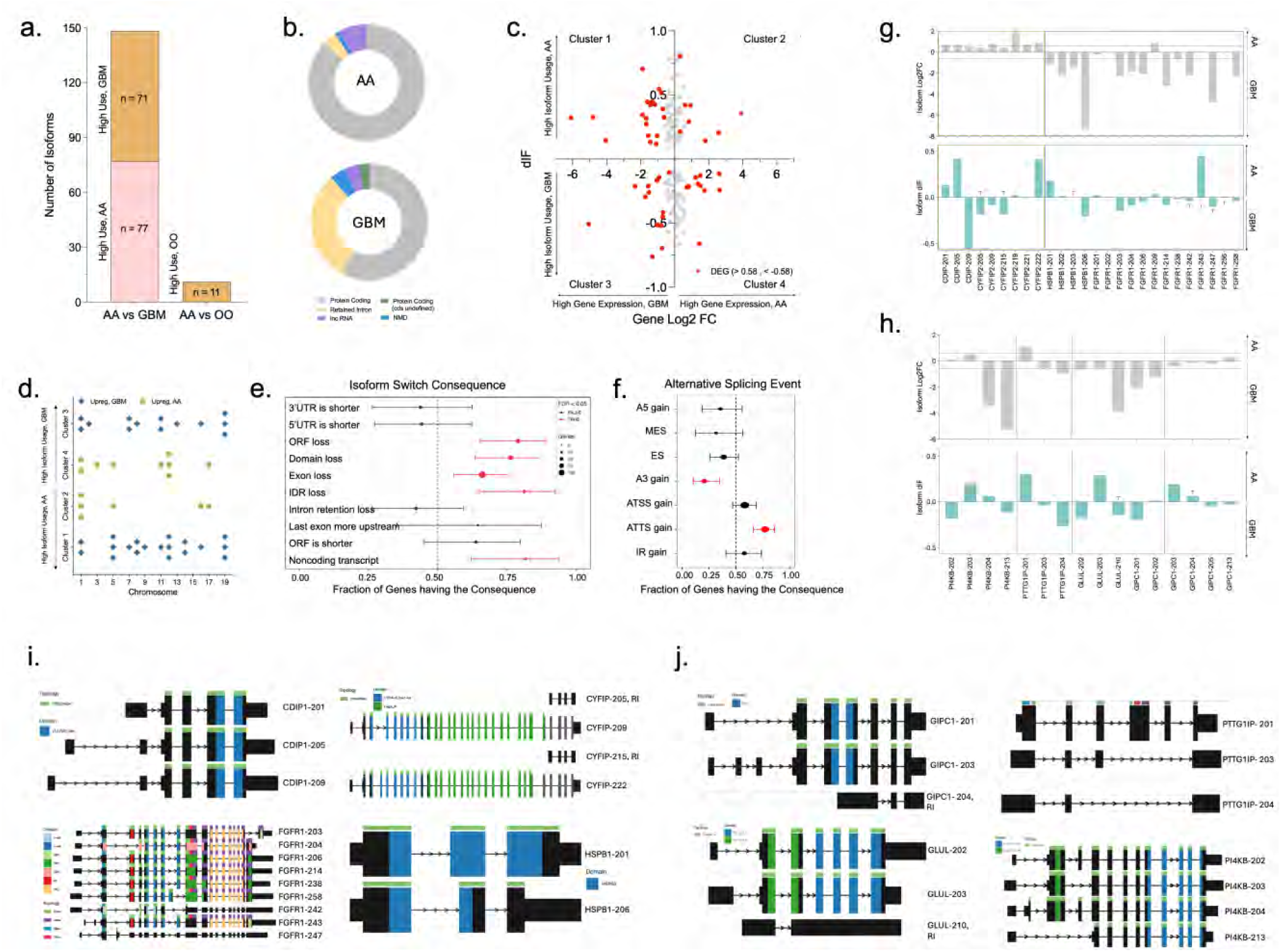
Differential isoform usage between Astrocytoma (AA) and Glioblastoma (GBM). **(a)** Stacked bar plot quantifying the number of high usage isoforms in AA vs GBM and AA vs OO. **(b)** Pie charts depicting the RNA biotype distribution among high usage isoforms in AA (n = 77, top panel) and GBM (n = 71, bottom panel). **(c)** Scatter plots showing the relationship between difference in isoform usage (difference in isoform fraction; dIF, y-axis) and gene level log2 fold change (x-axis) in AA vs GBM. Significant DEGs are highlighted in red. **(d)** Chromosomal distribution of upregulated genes in GBM (blue) and AA (green) linked to high or low isoform usage. **(e)** Consequence enrichment analysis identifying predicted structural consequences of isoform switches between AA and GBM. Y-axis: consequence, x-axis: fraction of genes having that consequence (95% confidence interval, CI). Dot size correlates with the number of genes with that consequence. **(f)** Alternative splicing event enrichment analysis in AA vs GBM. **(g)** Differential isoform expression (top, Log2 FC, top panel; gray) and usage (dIF, bottom panel; blue) in selected non-m6A modified upregulated genes in AA or GBM. **(h**) Differential isoform expression of m6A- modified transcripts with significant dIF usage in AA or GBM. Dotted line indicates a threshold for significance. **(i, j)** Isoform plots of genes depicted in **(g)** and **(h)**.

The integrated analysis of differential isoform usage (dIF) with gene level expression data (DeSeq) revealed four distinct expression-usage clusters (**Fig. 4c**): cluster 1 (high isoform usage in AA, high gene expression in GBM, n = 20), cluster 2 (high isoform usage and gene expression in AA, n = 8), cluster 3 (high isoform usage and gene expression in GBM, n = 14), cluster 4 (high isoform usage in GBM, high gene expression in AA, n = 10) (**Fig. 4c**). Overall, high isoform usage in AA (n = 20, Cluster 1) more frequently correlated with downregulated gene expression (upregulated; Cluster 2, n = 8) compared to GBM (**Fig. 4c**). Conversely, in GBM, high isoform usage in GBM (n = 14, Cluster 3) more frequently correlated with upregulated gene expression (downregulated; Cluster 4, n = 10) (**Fig. 4c**).

In AA vs. OO comparison, we identified 11 genes with significant isoform switches (n = 11) (**Supplementary Figure 8a**). All isoforms were used more in the OO subtype (**Supplementary Figure 8a**). We also identified statistically significant isoform switches in AA vs entry cortex and AA vs GBM comparisons (**Supplementary Figure 12**). Similar to the findings in tumor tissue, isoform expression and usage differences contributed to a difference at the gene level (**Supplementary Figure 12c-e**).

The isoform switch targets in each cluster exhibited distinctive patterns of chromosomal localization (**Fig. 4d**). Genes upregulated in AA (cluster 2, 4) frequently localized to the shorter chromosomes. In contrast, genes upregulated in GBM (cluster 1, 3), showed a more symmetric distribution across the chromosomes (**Fig. 4d**). Next, we sought to characterise the underlying structural features of isoform switches across the glioma subtypes (AA, GBM). Results from consequence enrichment analysis (*IsoformSwitchAnalyzeR*, **see Methods**) identified significant structural alterations in high usage isoforms in GBM (**Fig. 4e**): (i) complete open reading frame (ORF) loss, (ii) domain loss, (iii) exon loss, (iv) intrinsically disordered region (IDR) loss, and (v) non-coding isoform (**Fig. 4e**). Furthermore, splicing event analysis revealed that high usage isoforms in GBM were more likely to undergo alternative 3’ acceptor site (A3) gain and loss of alternative transcription start sites (ATTS), suggesting enhanced alternative splicing in GBM compared to AA (**Fig. 4f**). Importantly, statistically significant structural alterations and alternative splicing events were also identified in isoforms with differential usage in AA vs entry cortex and AA vs GBM (**Supplementary Figure 12a-b**).

Selected clinically relevant genes illustrate differences in isoform usage and gene expression: High usage of protein coding isoforms (CDIP1 - 205, CYFIP2-222; cluster 2) in AA correlated with higher gene expression compared to GBM (**Fig. 4g, 4i)**; High usage of NMD isoform (FGFR1-243) in AA correlated with lower gene expression compared to GBM (**Fig. 4g, 4i**). In GBM, high usage of protein-coding isoform HSPB1-206 correlated with higher gene expression compared to AA. (**Fig. 4g, 4i**). Further, we analyzed a subset of isoform-switched genes with confirmed m6A-modification (GIPC1, GLUL, PTTG1IP, PI4KB). These isoforms displayed differential usage and expression patterns, indicating potential transcriptomics regulation via m6A (**Fig. 4h, 4j**).

To explore the downstream functional implications of isoform switching we performed a gene ontology (GO) analysis (**see Methods, Supplementary Figure 8b-c**). In AA, the most enriched (ranked by fold enrichment) pathways included p53 mediated intrinsic apoptosis, TNF signaling, protein geranylgeranylation, tyrosine kinase signaling, and guanylyl cyclase signaling (**Supplementary Figure 8b-c**). In contrast, GBM associated enriched pathways were involved in oxygen carrier activity, inorganic phosphate transmembrane transport protein kinase C inhibitor activity, angiogenesis, hydrogen peroxide metabolism, and nitric oxide transport (**Supplementary Figure 8b-c**). Notably, pathways downregulated in AA were associated with inflammation, coagulation, epithelial to mesenchymal transition (EMT), and complement activation (**Supplementary Figure 8b-c**). Downregulated pathways in GBM were related to aldehyde-lyase activity, serine binding, postsynaptic calcium ion regulation, ubiquitin ligase activity, and hydroxymethyl- formyl- transferase activity (**Supplementary Figure 8b-c**).

### Long-read epitranscriptomic profiling in non-tumor, entry cortex tissue

We also analyzed *n=2* entry cortex tissue samples (samples 1, n=2 replicates and sample 2, n=3 replicates). We determined a higher prevalence of mapped reads in entry cortex 2 (30%) compared to entry cortex 1 (2%) (**Supplementary Figure 9a-b**), despite a similar RNA length distribution (**Supplementary Figure 9c**). Pooled analysis revealed approximately 1,000 m6A sites per transcript, over 500 genes (**Supplementary Figure 9d**) with most of the m6A modified transcripts falling under the protein coding category, followed by nonsense mediated decay (MMD) and retained intron (RI). Density plots revealed an increased m6A enrichment in the 3’UTR compared to gliomas **(Supplementary Figure 9e and Figure 1k)** and similarly to the glioma tumor RNA, no correlation was observed between m6A density and transcript length (**Supplementary Figure 9f**) with the primary kmer also being GGACT (**Supplementary Figure 9i**). Distribution analysis in entry cortex tissue demonstrated a high proportion of single m6A site methylated transcripts (**Supplementary Figure 9g-h**). Chromosomal distribution revealed an enrichment of m6A in chromosomes 19 (13%), 1 (11%), 16 (8%), and 11 (8%), similar to the glioma tumor RNA (**Supplementary Figure 9j).** Upregulated genes in gliomas included EEF1A1 (eukaryotic translation elongation factor 1 alpha 1), LTF (Lactotransferrin) and TUB1A1 (tubulin alpha 1a) while in the entry cortex tissue MT-TC (mitochondrially encoded tRNA-Cys) and RN7SL4P (RNA, 7SL, cytoplasmic 4) were upregulated compared to all gliomas subtypes (**Supplementary Figure 10a- c**). Integrative analysis of transcript methylation and gene expression across the groups revealed two genes upregulated in the entry cortex (YWHAH:14-3-3 protein eta, TMEM160:Transmembrane Protein 160) compared to all glioma subtypes with associated hypermethylated transcripts (**Supplementary Figure 10d-i**). Similarly, PUF60 (poly(U) binding splicing factor 60 protein) and TPT1 (Tumor Protein, Translationally-Controlled 1) were hypermethylated and upregulated in glioma vs. entry cortex (**Supplementary Figure 10d-i**). High usage isoforms in the entry cortex compared to glioma (AA, GBM) were shown to have a domain and exon loss (**Supplementary Figure 11a**). Statistically significant alternative splicing events in the high usage isoforms in entry cortex include ATTS (alternative transcription termination site) and ATSS (alternative transcription start site) gain (**Supplementary Figure 11b**). Consistent with previous observations in glioma (**Supplementary Figure 12**), up- and down- regulated gene expression in the entry cortex was shown to be attributed to the differential expression pattern of associated functional and non-functional isoforms (**Supplementary Figure 11c-d**).

### Transcript biotype composition and functional isoform burden across gliomas

In addition to mapping m6A modifications and isoform usage, we assessed the functional composition of transcripts across glioma subtypes. We defined functional isoforms as those annotated as protein-coding and non-functional isoforms as those classified as retained intron (RI), nonsense-mediated decay (NMD), or other non-coding biotypes. Although protein-coding isoforms predominated in all subtypes, their relative abundance differed: astrocytoma (AA) and oligodendroglioma (OO) samples showed higher proportions of protein-coding transcripts (71% and 70%, respectively), compared to glioblastoma (GBM, 67%) and entry cortex tissue (50–60%) (**Supplementary Table 9, Supplementary Figure 12a-b**).

However, when quantified at the gene level, only 8–16% of expressed protein-coding genes were composed almost entirely (≥75%) of functional isoforms, underscoring the substantial contribution of non-functional isoforms to the glioma transcriptome (**Supplementary Figure 12a-b)**. Notably, several amplified or highly expressed oncogenes, such as BCAN, HSPA5, and SPP1, displayed high proportions of non-functional isoforms (RI, NMD) in a subset of patients, despite overall gene upregulation (**Supplementary Figure 12a**). For instance, BCAN in OO and GBM harbored >70% non-coding isoforms in certain patients, while other astrocytoma cases showed as low as 5%, illustrating the inter-patient variability in isoform architecture (**Supplementary Figure 12a)**. Similarly, HSPA5 was amplified and upregulated in GBM yet exhibited >80% non-functional isoform expression in multiple samples (**Supplementary Figure 12a**).

Conversely, canonical glioma markers such as IDH1, MGMT, and BRAF consistently expressed protein-coding isoforms across all subtypes, suggesting tighter regulation of transcript functionality (**Supplementary Figure 12a**). These findings emphasize the importance of isoform-level quantification, as gene-level expression may not reflect true coding potential or translational competency in gliomas. Similarly, in glioma cell lines with knockdown writer (METTL3), reader (IGFBP2) and eraser (ALKBH5), we show differential expression of regulators and a shift from non-coding to protein-coding RNA isoforms, as illustrated in **Supplementary Figure 13**.

### m6a regulators and isoform-level associations reveal subtype-specific and clinically relevant RNA dynamics in glioma

To assess the clinical relevance of m6A modifications and upstream regulators, we analyzed the expression of 26 m6A associated proteins categorized as readers, writers, and erasers across the glioma subtypes (AA vs. GBM, OO vs. GBM, AA vs. OO) (**Fig. 5a**). Consistent with earlier findings of hypermethylation in IDH1 mutant gliomas, expression of m6A writers METTL3 and METTL14 was upregulated in the mutant group (**Fig. 5a**). Conversely, the demethylases ALKBH5 and FTO, which remove m6A sites, were upregulated in the IDH1 wild-type group, consistent with reduced methylation levels observed in GBM (**Fig. 5a**). Among the readers, we observed a distinct dichotomy in expression with YTHDF3 and YTHDDC2 upregulated in IDH1 mutant gliomas and YTHDF2 and IGF2BP2 elevated in GBM (**Fig. 5a**).

**Figure 5.**
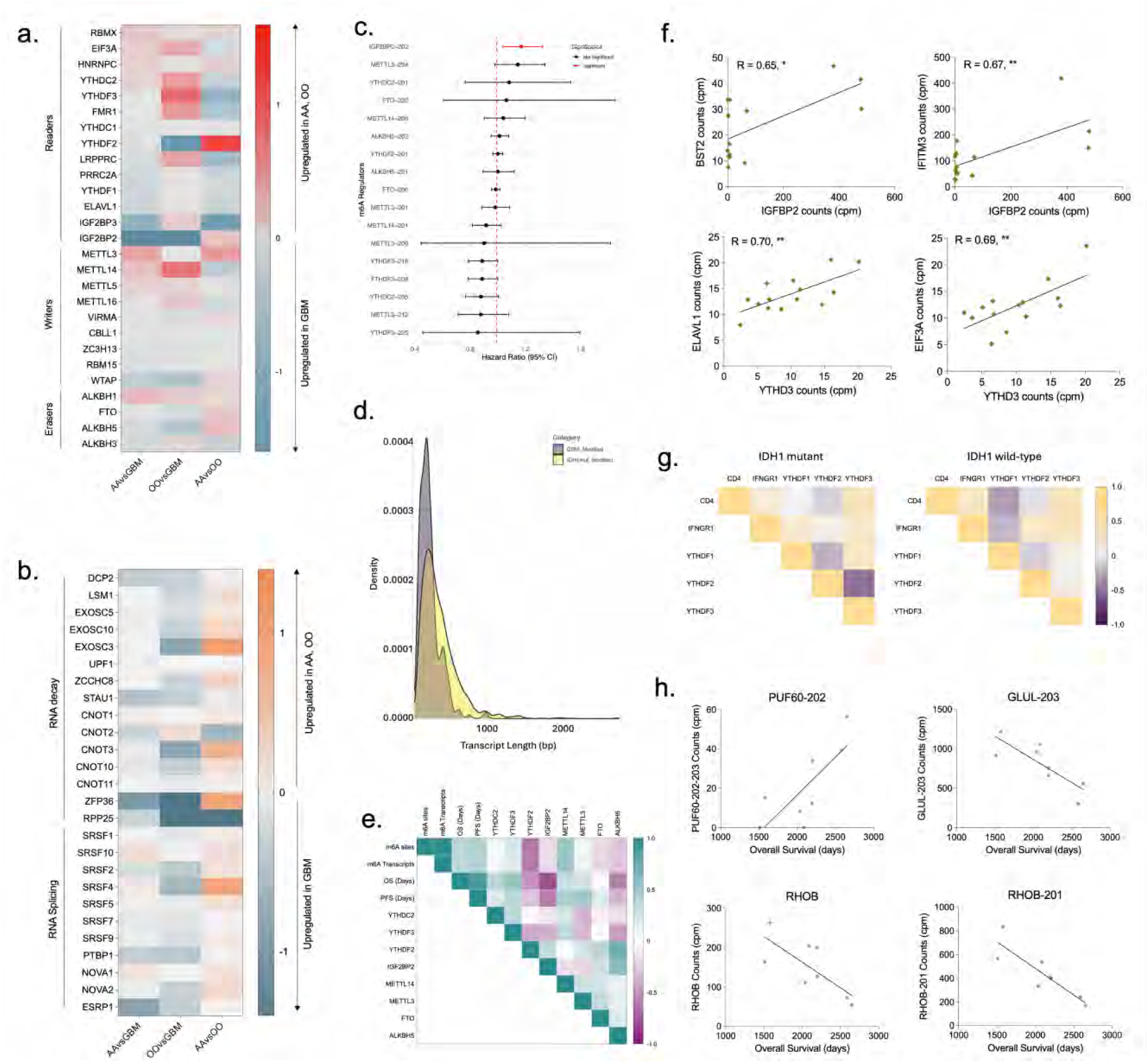
m6A regulators and transcriptomics consequences across glioma subtypes. Heatmap of differentially expressed (**a)** m6A regulators (readers, writers, erasers) and **(b)** RNA decay factors across three pairwise comparisons: AA vs GBM, OO vs, GBM, and AA vs. OO. **(c)** Correlation matrix of m6A regulator expression, m6A site/transcript abundance, and clinical outcomes (OS, PFS). **(d)** Forest plot of hazard ratios (HR, COX regression analysis) and 95% confidence intervals (CI) for isoforms of m6A regulators. Significant associations (p < 0.05%) are highlighted in red. **(e)** Density distribution comparing transcript lengths of m6A-modified RNAs in IDH1 mutant (yellow) and wild-type (purple) gliomas. **(f)** Gene expression correlation plots: IGF2BP2 with oncogenes BST2 (top, left) and IFITM3 (top, right); YTHDC3 with ELAVL1 (bottom, left) and EIF3A (bottom, right). **(g)** Correlation analysis (Pearson correlation, two-tailed test) between the m6A reader proteins and selected T Cell markers (CD4, IFNGR1) in IDH mutant (left) and wild-type (right) gliomas. **(h)** Correlation plots showing association of hypermethylated transcripts (PUF60-202, GLUL-203, RHOB-201) with overall survival in IDH1 mutant and IDH1 wild type gliomas.

YTHDF2 protein has previously been described in promoting RNA decay. We, therefore, next explored the subtype specific expression patterns of known markers of RNA degradation machinery (**Fig. 5b**). Overall, RNA decay factors including DCP2, EXOSC3, STAU1, CNOT3, and ZFP36 were all upregulated in GBM, particularly relative to OO (**Fig. 5b**). These findings indicate potentially enhanced RNA degradation in GBM. To support this, we examined the overall length distribution of m6A modified transcripts and found a shift toward higher prevalence of shorter, presumably degraded RNAs in GBM relative to IDH1 mutant gliomas (**Fig. 5d**).

Next, we evaluated whether the observed m6A modification levels and regulator expression differences correlated with clinical outcomes (**Fig. 5e**). Correlation matrix analysis revealed strong positive associations between m6A site/transcript abundance and both overall survival (OS) and progression free survival (PFS) (**Fig. 5e**). Among the regulators, positive prognostic markers included YTHDF3, YTHDC2, METTL3, and METTL14 (**Fig. 5e**). Negative prognostic factors included IGF2BP2, YTHDF2, ALKBH5, and FTO (**Fig, 5c**). To further identify the specific transcripts driving the m6A-survival associations, we performed a COX proportional hazard regression analysis on associated isoforms (**Fig. 5e**). Only IGF2BP2-202 showed a statistically significant negative correlation with OS (HR>1; p<0.05), implicating this isoform in glioma progression (**Fig. 5c**).

Potential pathogenic mechanisms and RNA regulatory effects of the m6A binding proteins (IGF2BP2, YTHDF3) were further examined using gene level correlations: IGF2BP2 was strongly correlated with BST2 and IFITM3, two known oncogenic genes (**Fig. 5f, top**), while YTHDF3 demonstrated positive correlation with ELAVL1 and EIF3A, supporting its reported role in translation enhancement (**Fig. 5f, bottom**).

To explore potential interaction between the m6A reader proteins and immune cell markers, gene expression correlation was performed (**Fig. 5g**). YTHDF3 was shown to demonstrate an overall positive correlation with CD4 and IFNGR1 in IDH1 mutant and wild-type gliomas. A negative correlation was noted for YTHDF1 in the wild-type group, but such pattern was not observed for the mutant group (**Fig. 5g**).

Finally, we examined how transcript specific m6A modifications correlate with survival (**Fig. 5h**). PUF60-202 was hypermethylated in AA and positively associated with survival (mut group), despite no no significance noted at the gene level (p = 0.144). We detected m6A modification in PUF60- 201, PUF60-202, PUF60-203, PUF60-204, and PUF60-208 (**Fig. 2c**). However, only PUF60-202 demonstrated a statistically significant correlation with overall survival. Similarly, we detected four distinct m6A modified GLUL isoforms (GLUL-201, GLUL-202, GLUL-203, GLUL-24) (**Fig. 2c**). Of these, GLUL-203 also showed hypermethylation in AA but was negatively associated with OS with statistical significance (**Fig. 5h**). Both RHOB gene and its transcript RHOB-201 were hypermethylated and negative prognostic in GBM (**Fig. 5h**). These findings illustrate the importance of isoform-level analysis in uncovering clinically meaningful m6A-mediated regulatory mechanisms that are otherwise obscured at the gene level.

## Discussion

IDH1 mutations are central to glioma classification, prognosis and therapy selection. These mutations produce 2-hydroxyglutarate (2-HG) which disrupts α-KG and its dependent enzymatic process, leading to epigenetic reprogramming^15^. While the role of 2-HG in promoting DNA hypermethylation is well established^15^, recent work has shown that it also inhibits RNA demethylases such as FTO, ALKBH5, resulting in reprogrammed m6A RNA methylation patterns^8,15,16^. Our study provides direct evidence supporting this mechanism by revealing subtype-specific m6A hypermethylation in mutant gliomas.

Across glioma subtypes, we observed that IDH-mutant astrocytomas and oligodendrogliomas exhibit higher levels of m6A-modified transcripts and significantly more m6A sites per isoform (AA- 2, OO-2; GBM-1) and m6A sites per gene (AA-20, OO-7, GBM-12) compared to IDH1 wild-type GBMs. These epitranscriptome differences coincide with distinct expression patterns of m6A regulatory machinery. Wild-type GBM preferentially expressed demethylase FTO and ALKBH5, consistent with prior reports^17^, while mutant tumors expressed higher levels of the methyltransferase METTL3 and METTL14. Notable, the expression of ALKBH5 varied between subtypes suggesting context-dependent 2HG sensitivity^18^

We also observed divergent expression of m6A reader proteins, with IGF2BGP2 and YTHDF2 enriched in GBMs and YTHDF3 elevated in IDH1-mutant gliomas. These patterns imply subtype-specific fates of methylated transcripts. YTHDF2 is a well-known mediator of m6A driven RNA decay^19^ and its high expression in GBM coincided with upregulation of RNA decay pathways, including decapping (DCP2^20^), the CCR-NOT deadenylase complex (CNOT3, CNOT10^20,21^), the 3’-5’ exosome complex (EXOSC3, EXOSC5, EXOSC10^22^), staufen-mediated decay (STAU1^23^), and ARE (AU rich element) mediated decay (ZFP36^24,25^). In contrast, YTHFD3 has been implicated in both translation and mRNA stabilization depending on the cellular context^19,26–28^. Its association with RNA-stabilizing proteins like eIF3A and ELAVL1 (HuR) in mutant gliomas suggests a potential role in transcript preservation and translation^29^.

We also found that the localization of m6A sites within transcripts influences expression outcomes. In astrocytomas, m6A enrichment in the 3’UTR of protein-coding transcripts was associated with increased expression, while in GBMs, m6A sites in coding sequences and 5’UTR were linked to elevated expression. These findings reflect broader observations that m6A positioning can direct diverse regulatory outcomes on RNA stability and translation^30,31^ and suggest that glioma subtypes may exploit distinct m6A codes to modulate gene expression. Intriguingly, we observed that retained intron isoforms, typically considered noncoding and unstable, were modified at the 5′ UTR in GBMs and correlated with high expression levels. This suggests that m6A may stabilize noncanonical isoforms in aggressive gliomas, potentially repurposing them for functional roles. These observations underscore a broader principle that m6A methylation may not only regulate gene expression at the level of canonical transcripts but also enable alternative isoforms to contribute to the transcriptome in subtype- and context-dependent ways.

These regulatory differences further extended to RNA isoform usage. Our transcript-level analysis uncovered widespread isoform switching between astrocytomas and GBMs, with over 148 isoform switches leading to significant expression changes in 52 genes; findings consistent with prior observations of splicing deregulation in high-grade gliomas^12,30,31^. GBMs preferentially, expressed noncoding isoforms such as retained introns and nonsense mediated decay transcripts, many of which were stabilized by m6A marks in the 5’UTR. These transcripts often exhibited structural alterations including shorter 5′ and 3′ UTRs, loss of open reading frames (ORFs), exon loss, and loss of functional domains or intrinsically disordered regions (IDRs), features known to affect transcript stability, translation, and protein function. Together with elevated expression of splicing factors in GBM, these findings support a model in which splicing dysregulation and m6A-mediated stabilization of noncanonical isoforms act in concert to shape the transcriptomic architecture of aggressive gliomas.

Pathway-level analysis further supports subtype-specific functional consequences of m6A regulations. IDH1 mutant gliomas showed enrichment for neurodevelopmental and Wnt signaling programs while GBMs exhibited activation of stress response, protein localization and adhesion pathways. These differences echo previous transcriptomic studies of glioma evolution and underscore the potential for m6A to fine–tune context specific cellular programs^32,33^.

Importantly, the expression of m6A machinery had prognostic implications, as previously reported^33^. Higher levels of METTL3, METTL14, YTHDF3 and YTHDC2 were associated with improved survival, whereas increased FTO, ALKBH5, IGF2BP2 and YTHDF2 correlated with worse outcomes. Notably, these associations were more pronounced at the isoform level: IGF2BP2-202, PUF60-202 and GLUL-203 were linked to poor survival despite no gene-level significance, highlighting the added resolution and clinical relevance of isoform-aware analysis^34^.

While our study offers a comprehensive view of the m6A epitranscriptome across glioma subtypes, several limitations remain. The sample size, particularly for oligodendrogliomas, was modest and requires validation in larger cohorts. Although nanopore direct RNA sequencing enables single-molecule methylation detection and isoform resolution, it may underrepresent low-abundance transcripts due to coverage constraints. Finally, while our analyses implicate specific m6A regulators and methylated isoforms in subtype-specific transcript regulation, causal relationships remain to be established through targeted perturbation studies.

In summary, we describe a glioma subtype–specific m6A landscape that influences RNA fate, isoform usage, and gene expression. These findings reveal distinct roles for m6A readers, writers, and erasers in shaping transcriptomic architecture and highlight the importance of isoform-level analysis in uncovering functional epitranscriptomic variation. Future studies using single-cell epitranscriptomics and CRISPR-based editing of m6A sites or regulators will be critical to elucidate cell type-specific effects and advance therapeutic targeting of RNA modification pathways in glioma.

## METHODS

### Study Population

The study population (n=14) included patients 18 years or older with histopathologically confirmed IDH1 mutant (n=8, Astrocytoma [AA]), n=6; Oligodendroglioma [OO], n=2) or IDH1 wild-type Glioblastoma (GBM, n=6) who underwent surgery at Massachusetts General Hospital (MGH) for biopsy or resection of a primary brain lesion. Exclusion criteria for the cohort included history of primary or metastatic cancers, active infectious disease (including SARS-CoV-2), and enrollment in clinical trials. In addition, we included non-tumor entry cortex tissue (n = 2) from patients undergoing tumor resection: metastatic melanoma (n = 1) and GBM (n = 1). Absence of EGFRvIII mutation in the entry cortex sample was confirmed via Digital Droplet PCR (ddPCR) using a previously developed assay^35^ (**Supplementary Fig. 9b**). Two replicates were sequenced for entry cortex patient 1 and three replicates for entry cortex patient 2 as the RNA yield was high and allowed for three replicates. All samples were collected with informed consent under Partners institutional review board (IRB)-approved protocol 2017P001581. Patient demographics are and clinical details are depicted in Supplementary Figure 2a.

### Tumor Tissue Processing

Tumor tissue aliquots are collected during neurosurgical resection or biopsy. Tumor tissue was micro dissected and suspended in RNAlater (Ambion) or flash-frozen, and stored at -80°C.

### Total RNA Isolation

Frozen tissue was thawed and lysed in 1-2 mL of ice-cold TriZol Reagent (ThermoFisher Scientific, Cambridge, MA, USA). Lysate was homogenized by passing through a 20-gauge RNAse-free needle 10 times. Total RNA was then extracted as per the manufacturer’s protocol and eluted in nuclease free water (Invitrogen). Both RNA quantity and quality were assessed for purity with Nanodrop One spectrophotometer (ThermoFisher Scientific, Cambridge, MA, USA). Agilent RNA 6000 pico kit was used with Agilent Technologies 2100 Bioanalyzer (Waldbronn, Germany) to determine the concentration and RIN (RNA Integrity Number) value of the samples.

### Ethanol precipitation

To remove potential contaminants and carry-over inhibitors, purification via ethanol precipitation was performed at multiple stages of the workflow: post extraction, post demethylation, and post poly(A)+ enrichment. To do this RNA was combined with 0.1 volume of 3 M, pH 5.2 sodium acetate and 3 volumes of ice-cold, 100% molecular biology grade ethanol (Sigma-Aldrich, St. Louis, MO). The ethanolic solution was stored at -20 ℃ overnight. Following this, RNA was recovered by centrifugation at 16,000g for 30 min at 4 ℃. The supernatant was carefully aspirated without disturbing the pellet. Subsequently, the pellet was washed with 0.5 ml of ice-cold, freshly prepared 70% ethanol. This was followed by centrifugation at maximum speed for 10 min at 4 ℃. The supernatant was removed, and the tube was left open at room temperature to ensure that last traces of fluid have evaporated. The pellet was then dissolved and resuspended in nuclease free water (Invitrogen).

### Poly(A)+ Isolation

Post enzymatic treatment with ALKBH5 or mock treatment, RNA samples were enriched for poly(A)+ species using the NEBNext® Poly(A) mRNA Magnetic Isolation Module (New England Biolabs,Ipswich, MA), according to manufacturer recommendations. All enrichment reactions were scaled up according to input RNA quantity, using 5 µg as upper limit for individual samples. Eluted poly(A)+ RNA was then assessed for quality and concentration by the RNA Pico mRNA Assay (Agilent).

### Library Preparation and Sequencing

Demethylated and mock treated poly(A)+ RNA from NEBNext poly(A) isolation module (New England Biolabs) was eluted according to manufacturer’s instructions, and then ethanol precipitated. RNA was pelleted and resuspended in 10 µL of nuclease free water (Invitrogen). 1 µL was used for RNA Pico mRNA Assay for quality check. The remaining 9 µL was used as input for library preparation. Libraries were prepared using the SQK-RNA002 kit (Oxford Nanopore Technologies) with selected modifications based on previous optimization runs: RTA and RMX ligation times were extended to 25 minutes, elution times were extended to 15 minutes, bead-binding times on Hula mixer were extended to 7 minutes, and Superscript IV (ThermoFisher Scientific) was used instead of Superscript III (ThermoFisher Scientific). As such, thermocycling conditions were modified, and the RTA adapted RNA was reverse transcribed at 53°C for 50 minutes, with reaction inactivation at 80°C for 10 minutes, before holding at 4°C. Following library preparation, demethylated and mock treated poly(A)+ RNA samples were sequenced on a MinION sequencer using R9 flow cells (Oxford Nanopore Technologies) for 24 hours, or until refuel of flow cell resulted in a lack of reads. Live basecalling (fast) was used to monitor Q-score (Qfilter ≥ 5) and translocation speed for the purposes of refueling.

## Statistical Analysis

### Raw Sequencing throughput, QC, Genome Alignment, and Quantification

Total RNA extracted from each patient tissue sample was split into two aliquots, demethylated along with a mock control, and sequenced in parallel. Raw FAST5 files were compiled for each sequencing run, and pass reads (qscore>7) were basecalled using Dorado (0.5.0+0d932c0) using the model rna002_70ps_fast@v3. The resulting FASTQ files were processed using nanoseq^36^(3.1.0) (https://github.com/nf-core/nanoseq/tree/dev), an analysis pipeline for Direct RNA Sequencing Data. It comprises raw read QC, alignment, and quantification.

During the first part of nanoseq, QC metrics from raw reads were generated using Nanoplot. Next, reads were aligned to the human (GRCh38) genome using minimap2 (2.17-r941). Post alignment, SAM files were converted to sorted BAM files using samtools (1.16.1) and mapping metrics were presented using MultiQC (1.11). Finally, nanoseq utilized bambu (3.0.8) to quantify human genome alignments and generate gene counts and normalized abundances. The resulting raw count data were imported into R for downstream analyses.

### Enzymatic Demethylation

Total RNA extracted from each tumor tissue was equally split into two aliquots for subsequent demethylation or mock treatment. Given the variability in RNA yield from each tissue sample, up to 200 μg of RNA was either demethylated or mock treated with active recombinant FTO/ALKBH5 protein (Abcam, Cambridge) at a 1:0.3 molar ratio in a 500 µL reaction, as previously described by Zheng et. al, in 50 mM HEPES (Sigma-Aldrich, St. Louis, MO), 100 µM 2-oxoglutarate (Sigma-Aldrich, St. Louis, MO), 100 µM ascorbate (Sigma-Aldrich, St. Louis, MO), 50 µM Ammonium (II) Iron Sulfate (Sigma-Aldrich, St. Louis, MO), 1 mM TCEP (Sigma-Aldrich, St. Louis, MO), and 50 U of RNAse-Inhibitor (ThermoFisher Scientific, Cambridge, MA, USA). Care was taken to avoid introduction of RNAses, and all solutions were prepared in nuclease free water (Ambion). RNA was ethanol precipitated and eluted in 100 µL of nuclease free water.

### Quality Control Metrics for ALKBH5 Treated and Untreated

Initially the study design was based on MasterofPores for paired m6A modification prediction. However, while the analysis was ongoing, m6Anet was developed which accurate prediction of m6A modifications without the need for a treated RNA sample. Our QC analysis (**Supplementary Figure 2-3**) the treated and untreated samples were similar in terms of read depth and predicted m6A sites. Principal component analysis (PCA) was performed to visualize clustering patterns in gene expression data from ALKBH5-treated and untreated RNA samples in AA, OO, and GBM cohorts. Venn diagrams were used to assess the overlap and uniqueness of detected genes between treated and untreated samples across individual patients. The distribution of detected m6A sites was compared between ALKBH5-treated and untreated RNA samples, and m6A site distributions were examined across individual patients to assess consistency and variability in modification profiles. Due to the high degree of similarity observed between treated and untreated samples, they were subsequently combined and treated as biological replicates in downstream analyses.

### Differential expression and isoform usage analysis

Differential gene and isoform analysis was performed using *DESeq2*^37^ (1.45.3) in R. Prior to analysis, for each patient with ALKB-treated and untreated conditions, raw counts were aggregated across conditions. Lowly expressed genes were filtered out by removing genes with less than 10 counts across all samples. The remaining counts were normalized using DESeq’s internal size factor estimation. Log2 fold changes and adjusted p-values (Benjamini-Hochberg) were calculated, with an adjusted p-value (FDR) threshold of 0.05. Differential isoform usage analysis was performed in R using IsoformSwitchAnalyzeR^38^ (2.5.0). The aggregated isoform counts and abundances were input along with the human (GRCh38.p14) annotation and transcriptome files. Single isoform genes were filtered out during the *preFilter()* step since these genes cannot exhibit changes in isoform usage. Statistical analysis was performed with *isoformSwitchTestSatuRn()*. We required a difference in isoform proportions between classification groups of >0.2 and an FDR-adjusted p-value of <0.05 for significance. The functional consequences of the identified isoform switches were generated using the analyzeSwitchConsequences() function. This analysis identified changes in key functional domains such as coding potential, exon loss, domain loss, and isoform length. Alternative splicing analysis was performed using the extractSplicingEnrichment() function, which identified and assessed the significance of specific events such as alternative 3’ acceptor site (A3) gain or loss.

### Transcriptome-wide m6A modification sites

We used m6Anet^14^ (2.1.0), a machine learning-based tool, to detect m6A sites in DRACH motifs from our direct RNA reads in all samples. Input data included raw nanopore FAST5 reads, which were aligned to the reference genome using minimap2. After alignment, m6ANet was employed to predict m6A modification sites based on sequence context and nanopore signal patterns. As m6anet supports pooling over replicates, treated and untreated samples for each patient were pooled prior to the inference step. The output of m6Anet provided predicted m6A sites with probability that the site is modified, the transcript position of the site, the 5-mer motif of the site, and the estimated percentage of reads in the site that is modified. For our analysis, we defined high confidence m6A sites as those with a probability modified value greater than or equal to 0.9. Group-level m6A sites were defined as those detected in at least three patients within the AA and GBM groups, and all detected sites within the OO group.

### Distribution of modified sites

To analyze the distribution of m6A-modified sites across transcript regions, custom R scripts were used to calculate the lengths of the 5’ UTR, CDS, and 3’ UTR regions. For each transcript, the coding sequence (CDS) length was determined by summing exon lengths, while the UTR lengths were calculated based on start and end coordinates. Transcripts with missing UTR annotations were assigned a length of zero for the missing regions. To visualize the distribution of m6A-modified sites across these regions, a custom function was implemented in Python. The transcript position of each modification site was compared with the UTR and CDS regions to determine if the site was located within the 5’ UTR, CDS, or 3’ UTR. The relative position within each region was calculated, and kernel density estimation (KDE) plots were generated to visualize the relative density of m6A sites across these transcript regions. Modifications grouped by transcript biotypes (protein-coding, nonsense mediated decay, retained intron) and methylation status were visualized.

### Identification of hyper- and hypo- m6A methylated Transcripts

To identify commonly modified sites within each tumor classification, we first filtered for sites present in at least three patients with AA, at least three patients with GBM, and at least one patient with Oligoastrocytoma (OO). For these commonly identified sites within each group, we then identified transcripts containing sites common to all three classifications (AA, GBM, and OO). For these commonly modified transcripts, we calculated the average modification ratio for each site within each classification. Finally, to calculate the weighted modification ratio for these commonly modified transcripts, we summed the modification ratios of all sites within each transcript and divided by the transcript length. This approach ensured that transcripts with more modified sites contributed proportionally while preventing length bias, allowing for a more accurate comparison of modification levels across transcripts of varying lengths. To identify hypermethylated and hypomethylated transcripts, we calculated the log2 fold change of the weighted modification ratio for each pairwise comparison (AA vs. OO, OO vs. GBM, AA vs. GBM). A positive log2FC indicated that a transcript was hypermethylated in AA (vs. GBM), AA (vs. OO), or OO (vs. GBM), while a negative log2FC indicated that a transcript was hypermethylated in GBM (vs. AA), OO (vs. AA), or GBM (vs. OO).

### Functional Analysis of Isoform Switching and hyper- and hypo- m6A methylated Genes

Functional analysis was performed using ClusterProfiler^39^ (4.14.6) for all Gene ontologies (Biological Process, Molecular Function, and Cellular Component) and KEGG pathways. The enrichGO function was applied for genes that were upregulated and had m6A modifications in each tumor classification.

### m6A Regulators Correlation with Survival Outcomes

Clinical metadata and expression data for m6A regulators included overall survival time (in days), vital status (coded as 1 = deceased, 0 = censored), IDH1 Status, and normalized transcript expression values. To evaluate the prognostic significance of individual m6A regulators, univariate Cox proportional hazards models were fit for each gene using the coxph() function from the surviva^l40^ package in R. For each transcript, a model of the form *Surv(time, status) ∼ transcript expression* was fit to estimate the hazard ratio (HR), 95% confidence interval, and p-value. Variables not converging were excluded. Hazard ratios greater than 1 were interpreted as indicating higher risk associated with increased expression, while values less than 1 suggested a better prognosis with increased expression. Transcripts with a p-value less than 0.05 were considered statistically significant. A forest plot was generated using ggplot2 to visualize the results, with error bars representing 95% confidence intervals and color coding to indicate statistical significance.

### Gene Fusion Detection

Gene fusions were quantified using JAFFA (https://github.com/Oshlack/JAFFA/wiki). FASTQ files were used as input and the JAFFA.groovy script was used with the accurate ONT aligner minimap2 to maximise sensitivity for fusion detection. The output includes genes involved in the fusion event, the position of the fusion, the number of reads, and classification (HighConfidence, MediumConfidence, LowConfidence, and PotentialTransSplicing), and whether or not the fusion is known.

### Institutional review board statement

Our studies were conducted in accordance with principles for human experimentation as defined in the U.S. Common Rule and were approved by the Human Investigational Review Board of each study center under Partners institutional review board (IRB)-approved protocol number 2017P001581. All healthy control subjects were screened for pertinent oncologic and neurologic medical histories. Individuals with a history of cancer, neurological disorders, and infectious diseases were excluded from the study.

### Informed consent statement

All samples were collected with written informed consent after the patient was advised of the potential risks and benefits, as well as the investigational nature of the study.

## Data Availability

All data produced in the present study are available upon reasonable request to the authors

## Data availability

The sequencing and m6A modification data generated in this study are provided in the Supplementary Information and Source Data file. The raw nanopore sequencing data and m6Anet output are not publicly available but will be available upon request from the corresponding author. Source data are provided with this paper.

## Acknowledgements

The authors would like to thank all the MGH Neurosurgery clinicians and staff who assisted with the collection of samples. We are also deeply appreciative to the patients and their families for participating in the study. This work is supported by grants R01 CA239078, CA237500, CA291826 (BSC, LB). MGN Transformative Scholar (LB), Rappaport Scholar (LB). The funding sources had no role in the writing of the manuscript or the decision to submit the manuscript for publication. The authors have not been paid to write this article by any entity. The corresponding author has full access and assumes final responsibility for the decision to submit for publication.

## Contributions

S.M.B., H.L., K.M., S.M.K. L.B., Data curation, formal analysis, validation, investigation, visualization, writing of original draft, writing, review and editing. A.K.E., T.H., Experimental work, analysis, review and editing; E.E., A.K., Sample collection, processing, clinical correlates, data analysis: D.P.C., G.P.D., J.J.M., B.D.C., A.S.P., B.S.C., Formal analysis, methodology, review and editing. L.B., Conceptualization, resources, data curation, software, formal analysis, supervision, funding acquisition, validation, investigation, visualization, methodology, writing—original draft, project administration, writing—review and editing.

