## supplementary information for "IDH1-dependent m6A methylation defines transcriptomic heterogeneity in glioma"

#### Supplementary Figure 1

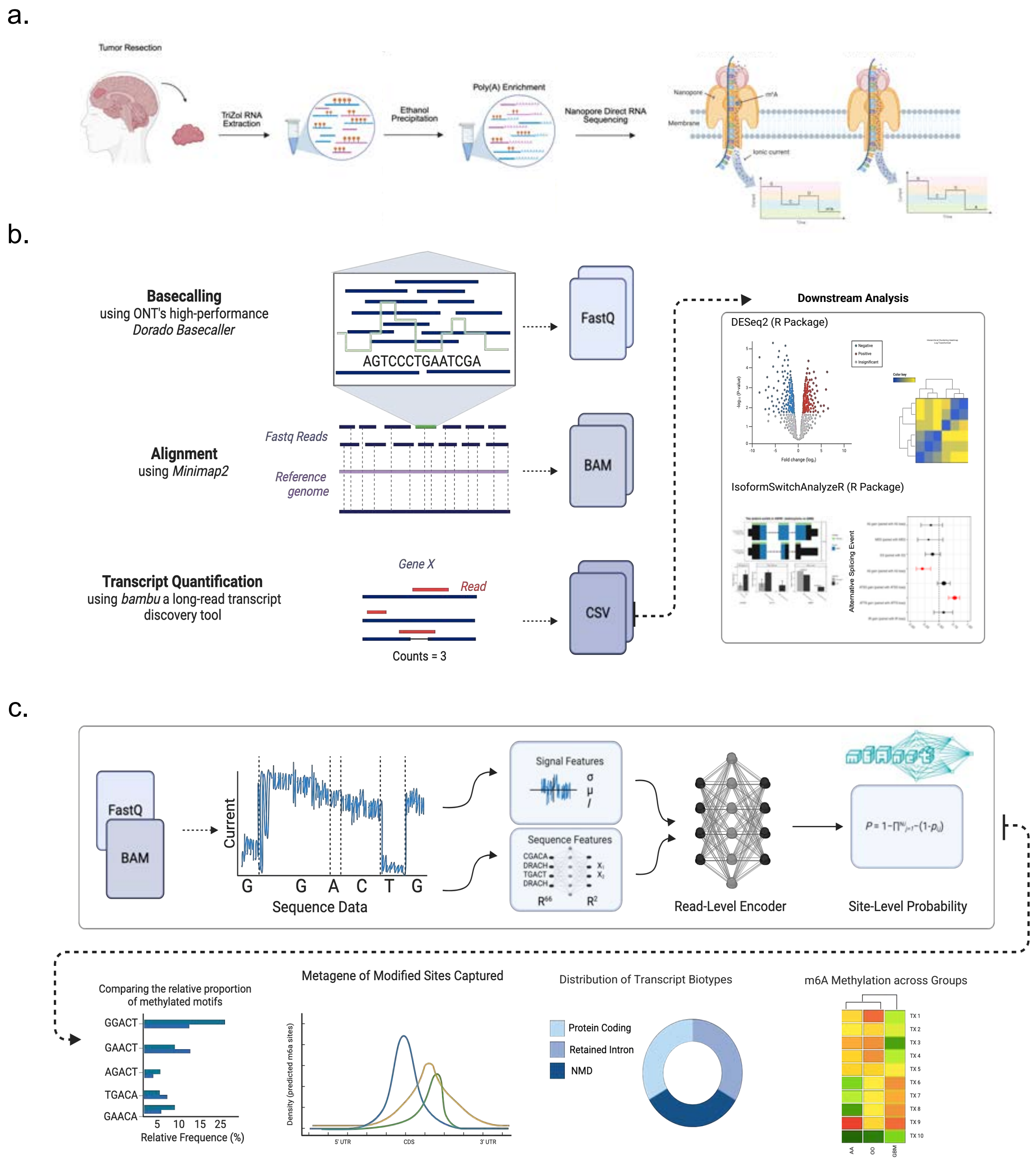

**Supplementary Figure 1. Overview of direct RNA sequencing and m6A analysis pipeline** (a) Experimental Workflow. Total RNA from glioma tissue was poly(A) enriched and sequenced using Oxford Nanopore direct RNA sequencing. (b) Post-sequencing analysis. Reads were base-called (Dorado, aligned to the human genome (Minimap2), and quantified using a long-read transcript tool (Bambu). Resulting data (FastQ, BAM, CSV) were used for differential gene and transcript expression (DESeq2) and isoform usage analysis (IsoformSwitchAnalyzeR). (c) m6A modification analysis. Using m6anet, signal and sequence features were extracted to infer site-level m6A probabilities using a neural network model. Outputs include motif enrichment, metagene profiles, biotype distributions, and group-level m6A comparisons.

#### Supplementary Figure 2

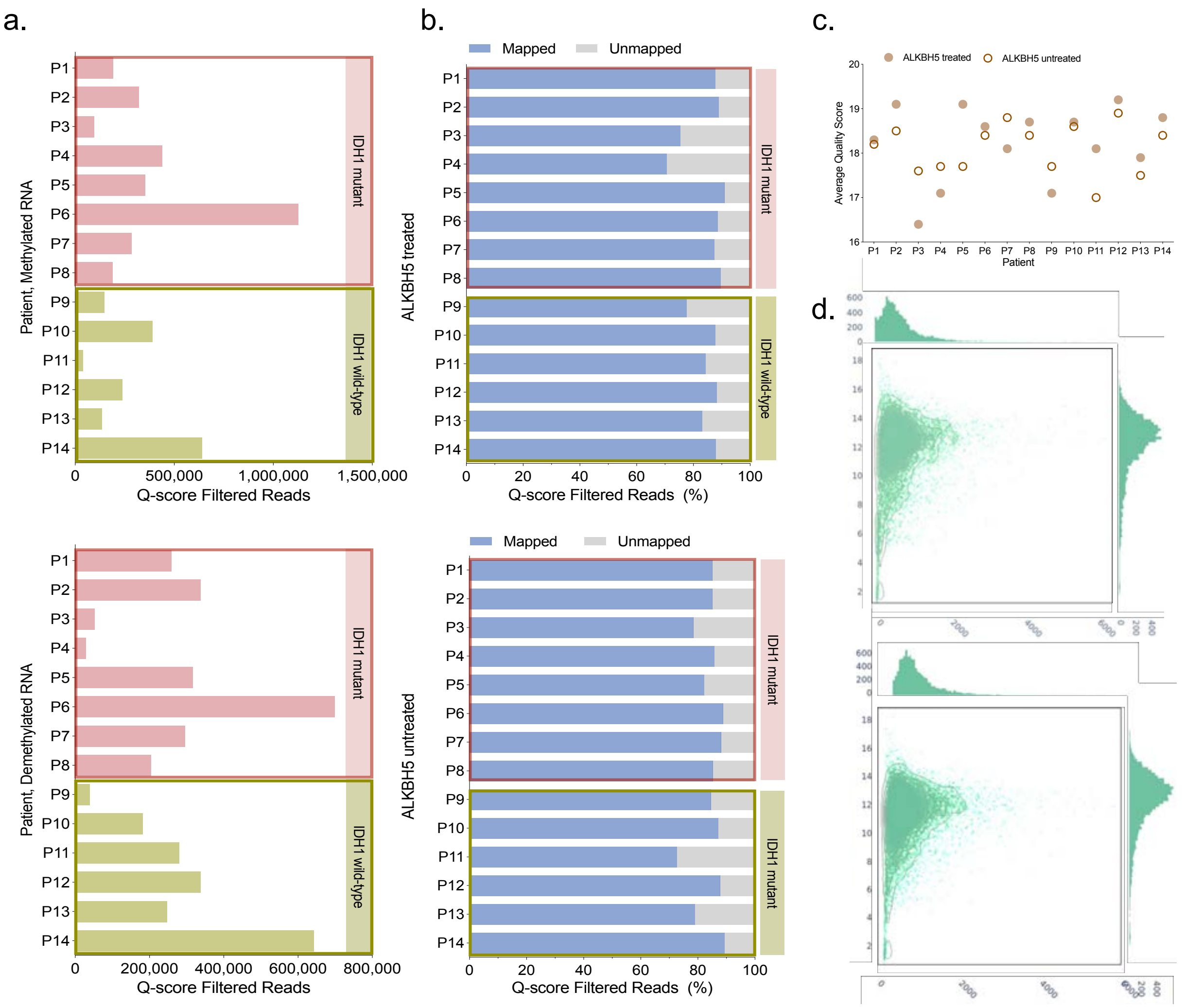

**Supplementary Figure 2. Comparison of sequencing output in ALKBH5 treated and untreated RNA.**

(a) Bar graphs depicting the number of Q score filtered reads in ALKBH5 treated (top) and untreated (bottom) RNA sequenced from individual patients. (b) Distribution of mapped and unmapped reads during alignment for ALKBH5 treated (top) and untreated (bottom) samples. (c) Dot plot comparing the the average quality score of the sequencing output from treated and untreated RNA in individual patients. (d) Quality control (QC) Nanoplots plotting the log transformed read lengths against average read quality (using a kernel density estimate) in ALKBH5 treated (top) and untreated (bottom) samples.

**Supplementary Figure 3**

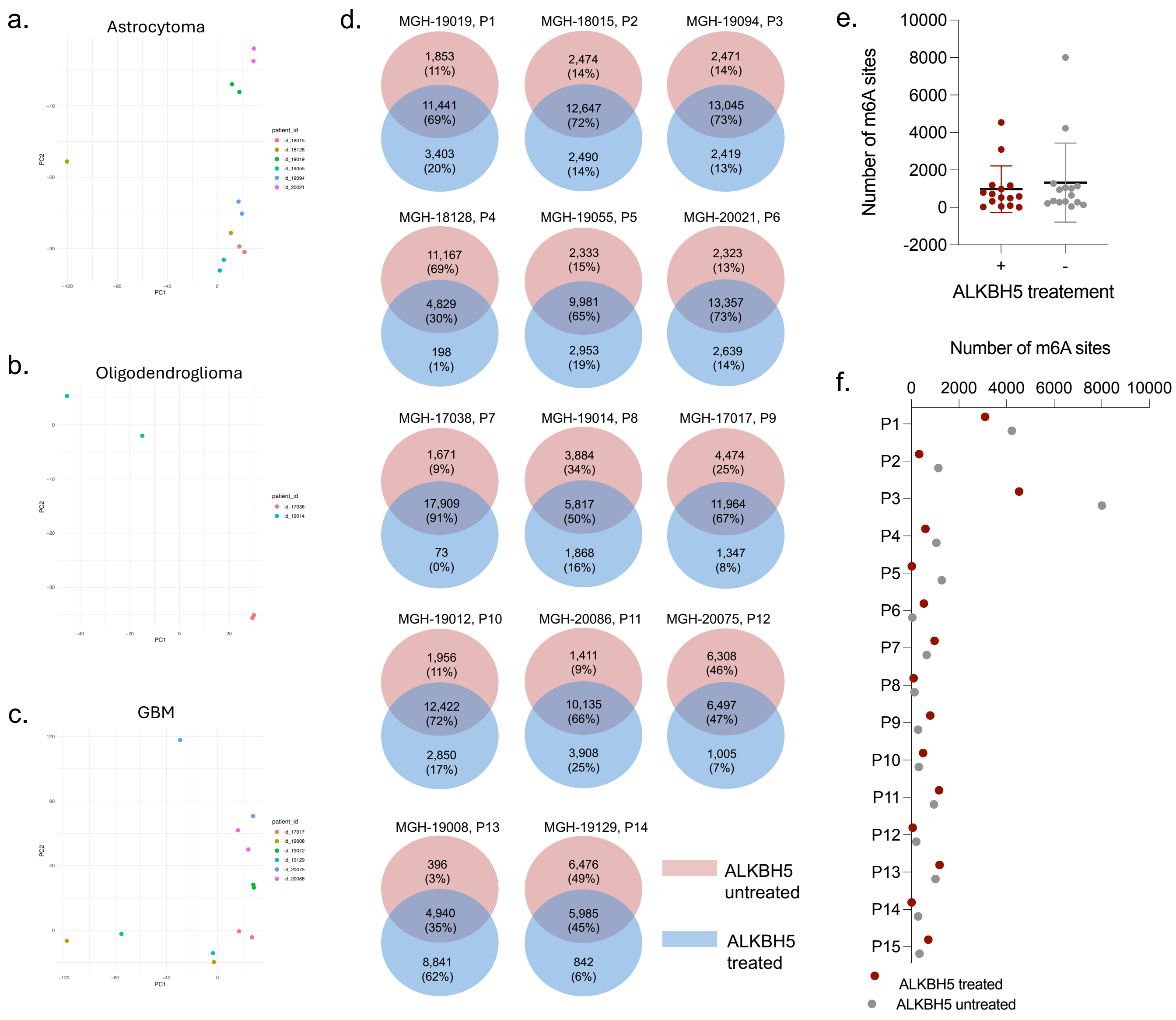

**Supplementary Figure 3. Comparison of gene expression and m6A site detection with ALKBH5 treatment**. (a-c) PCA plots highlighting the clusters of gene expression data for ALKBH5 treated and untreated RNA in AA (a), OO (b), and GBM (c). (d) Venn diagrams demonstrating the number and percentage of common and unique genes detected in ALKBH5 treated and untreated RNA across individual patients. (e) Distribution of detected high confidence (probability modified > 0.9) m6A sites between ALKBH5 treated (red) and untreated RNA (gray) with annotated median and SD. (f) Distribution of detected high confidence) m6A sites between ALKBH5 treated (red) and untreated RNA (gray) across individual patients.

#### Supplementary Figure 4

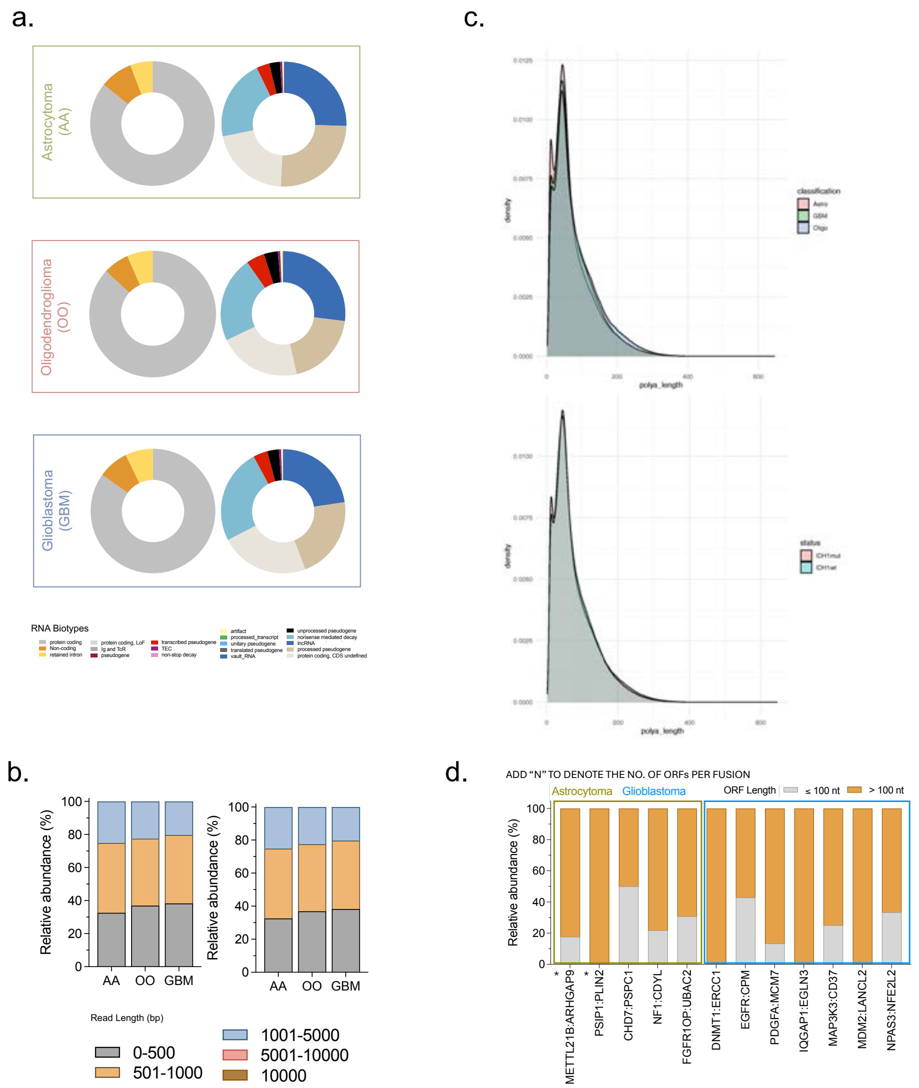

**Supplementary Figure 4. Distributions of key transcriptomic features across glioma subtypes.**

(a) Pie charts showing the distribution of RNA biotypes across glioma subtypes. The left panel displays the proportions of protein-coding, retained intron, and non-coding transcripts. The right panel further breaks down the composition of the non-coding transcript category. (b) Stacked bar chart showing the relative abundance (%) of mapped read lengths of overall (left) and full-length (right) annotated transcripts across subtypes. (c) Poly-a tail density distribution across (top) AA, OO, and GBM and (bottom) IDH1 mutant and IDH1 wild-type. (d) Relative abundance and ORF length distribution of detected fusion transcripts in AA and GBM. Asterisks denotes high confidence fusion transcripts.

#### Supplementary Figure 5

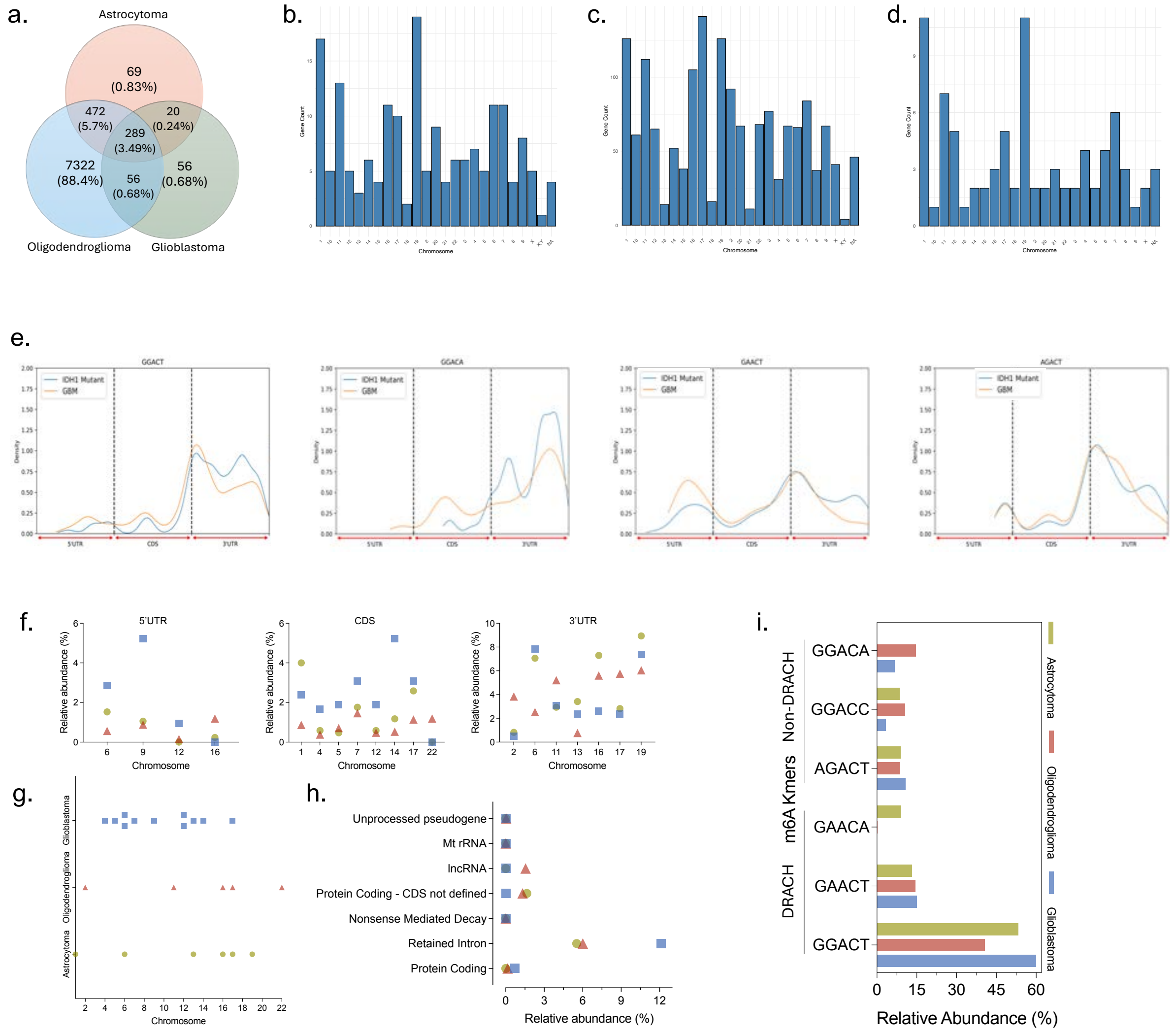

**Supplementary Figure 5. Transcriptomic analysis of detected m6A modified sites.** (a) Venn Diagram quantifying the shared vs. unique m6A sites across the three glioma subtypes. (b-d) Chromosomal distribution of the filtered m6A sites (probability of modification ≥ 0.90), highlighting the frequency of modified sites per chromosome across three glioma subtypes: (b) Astrocytoma, (c) Oligodendroglioma, (d) Glioblastoma. (e) Density plots representing the distribution of the top 5 m6A kmers along the transcript regions (5' UTR, CDS, and 3' UTR) across the glioma subtypes. (f) Scatter plots demonstrating the spatial relative distribution of m6A modified genes in Astrocytoma (green circle), Oligodendroglioma (red triangle) and Glioblastoma (blue square) across the distinct transcript regions: 5'UTR (left), CDS (middle), and 3'UTR (right). (g) Scatter plot demonstrating the chromosomal distribution of the m6A modified genes across the glioma subtypes. Only the targets where the prevalence of m6A sites differs by ≥ 0.5% between any two groups are plotted. (h) A comparative RNA biotype distribution analysis of the m6A modified transcripts in the 5'UTR region using the scatter plot. (i) Distribution of the most prevalent m6A motif (GGACT), and the top 5 DRACH and non-DRACH Kmers across the glioma subtypes.

#### Supplementary Figure 6

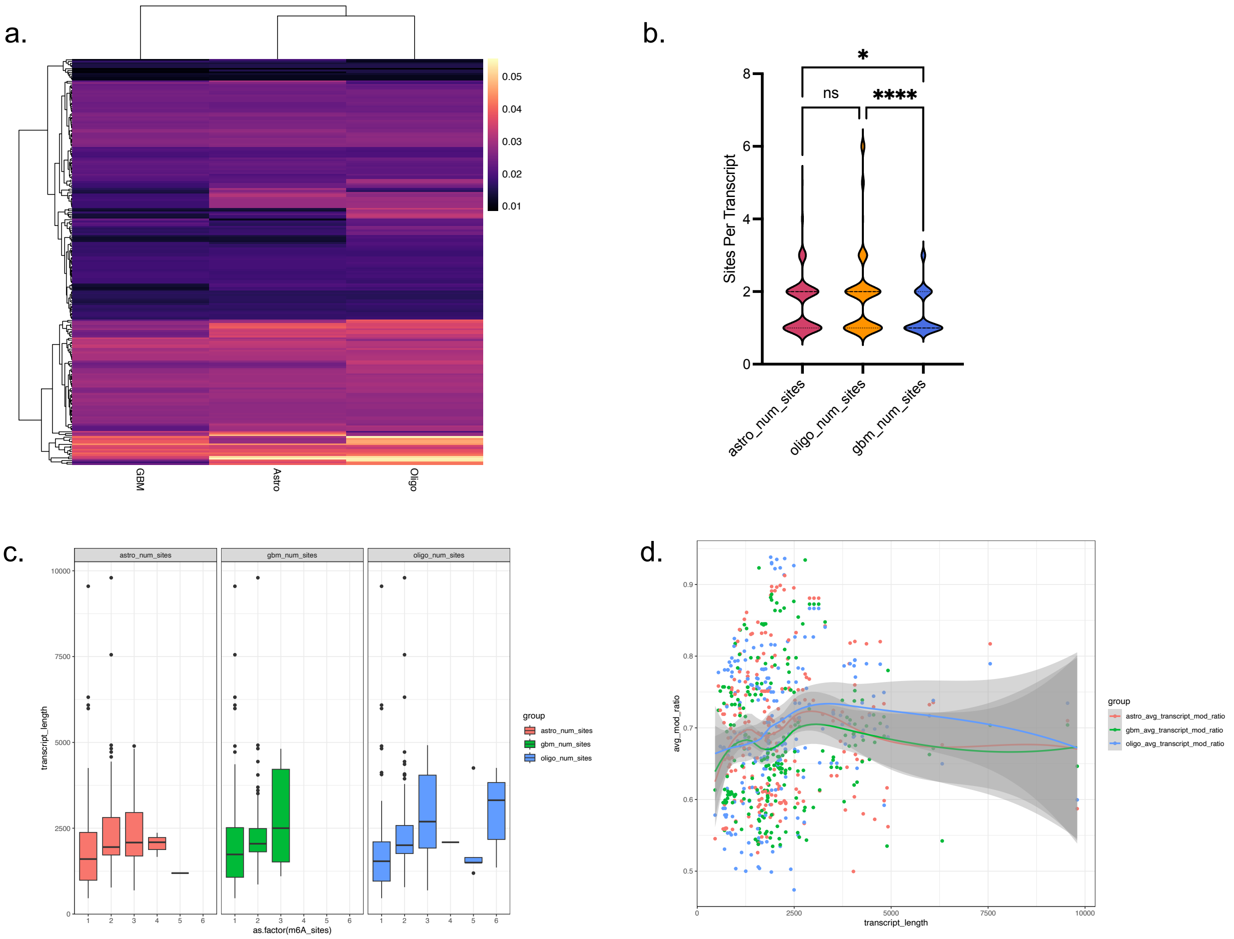

**Supplementary Figure 6. Commonly modified transcripts across the glioma subtypes demonstrate variation in m6A levels.**

(a) Heatmap illustrating the distribution of the calculated weighted modification ratio for commonly m6A-methylated transcripts across all classifications. A square root transformation was applied to enhance visualization of differences in modification levels. Black indicates transcripts that were not modified. (b) Violin plots depicting the distribution of sites per transcript for the commonly methylated transcripts. A significant difference was observed among AA (red), OO (orange), and GBM samples (blue) (Kruskal-Wallis test, $p < .05$). (c) Boxplots showing the distribution of transcript length (y-axis) across different numbers of m6A sites per transcript (x-axis) for AA (red), GBM cells (green), and OO (blue). (d) Distribution of average modification ratio per site (y-axis) across transcript length (x-axis) per classification.

### Supplementary Figure 7

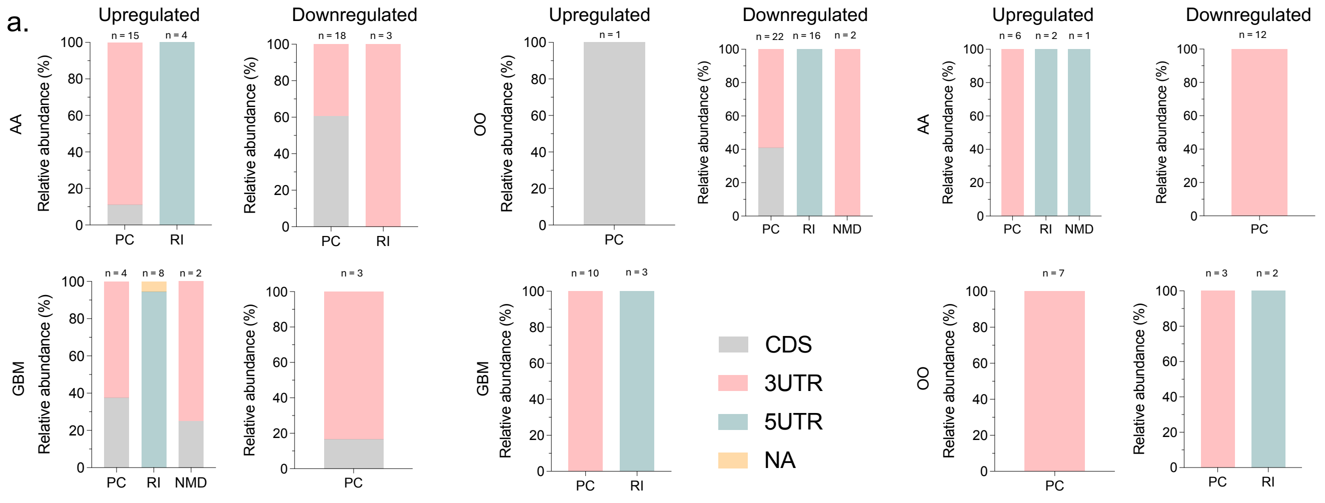

b.

| Hypermethylated Group \ Upregulated Gene Group | AA | GBM |
|---|---|---|
| AA | EEF1A1 (16)<br>LDHB (3) | ACTB (6)<br>AP2S1 (5)<br>LGALS3BP (4)<br>ATP6V0E1 (2)<br>S100B (2)<br>CEBPD (1)<br>CENPB (1) |
| GBM | BCAN (1)<br>EEF1A1 (1)<br>EEF2 (1) | HSPA5 (6)<br>ACTB (4)<br>APOD (2)<br>TMSB10 (1)<br>SPP1 (1) |

c.

| Hypermethylated Group \ Upregulated Gene Group | OO | GBM |
|---|---|---|
| OO | | APS21 (5)<br>HSPA5 (5)<br>TUBB2A (5)<br>LGALS3BP (4)<br>DKK3 (3)<br>S100B (2)<br>CENPB (1) |
| GBM | BCAN (1) | DYNLL1 (4)<br>C1orf122 (3)<br>CEBPD (1)<br>TMSB10 (1)<br>HSPA5 (1) |

d.

| Hypermethylated Group \ Upregulated Gene Group | AA | OO |
|---|---|---|
| AA | TUBB2A (4)<br>C1orf122 (3)<br>CEBPD (1) | C1QA (8)<br>HTRA1 (1) |
| OO | DKK3 (3)<br>TUBB2A (1) | HTRA1 (2) |

**Supplementary Figure 7. Biotype and region distribution with upregulated and hypermethylated genes in glioma.**
(a) Integrated biotype and transcript region distribution for different clusters of upregulated and hypermethylated gene groups per group in comparisons across AA vs. GBM (left), OO vs. GBM (middle), AA vs. OO (right). PC represents protein coding, RI represents retained intron, and NMD represents Nonsense-mediated decay transcripts. (b-d) Upregulated genes and hypermethylated gene groups in different sample comparisons (AA vs. GBM, left; OO vs. GBM, middle; AA vs. OO, right), with numbers indicating the number of transcripts for each gene within each overlapping category.

#### Supplementary Figure 8

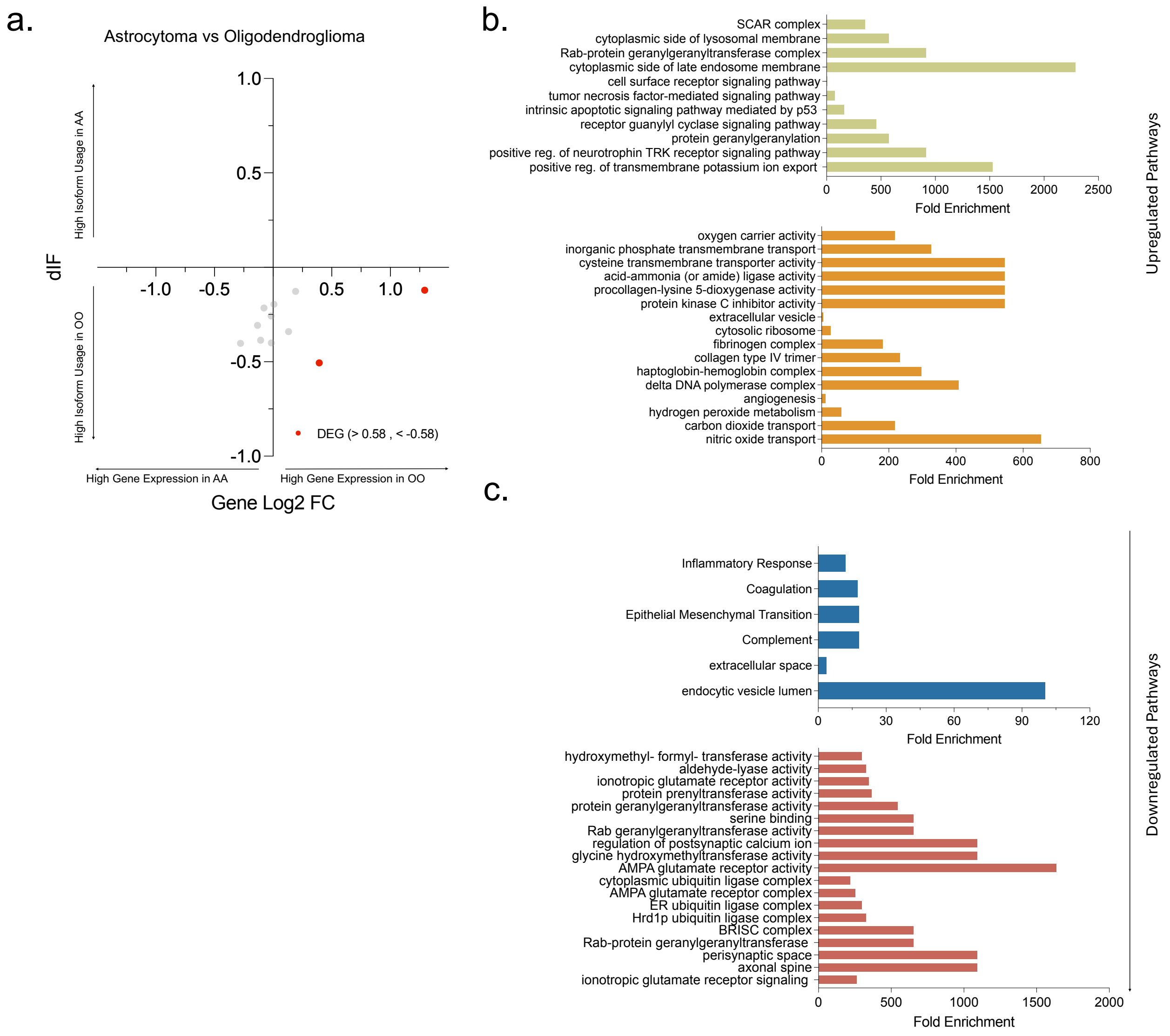

**Supplementary Figure 8. Isoform Switching Analysis with Gene Expression and Functional Pathways** (a) Scatter plot of differences in isoform fraction (dIF, y-axis) and differential gene expression (Log2 FC, x-axis) for AA vs OO comparison, with significant points highlighted in red. (b) Gene ontology (GO) analysis of significantly upregulated genes in AA (top) and GBM (bottom) to identify upregulated pathways. Fold enrichment plotted for significantly enriched pathways. (c) GO analysis of significantly downregulated genes in AA (top) and GBM (bottom) to identify downregulated pathways. Fold enrichment plotted for significantly enriched pathways.

#### Supplementary Figure 9

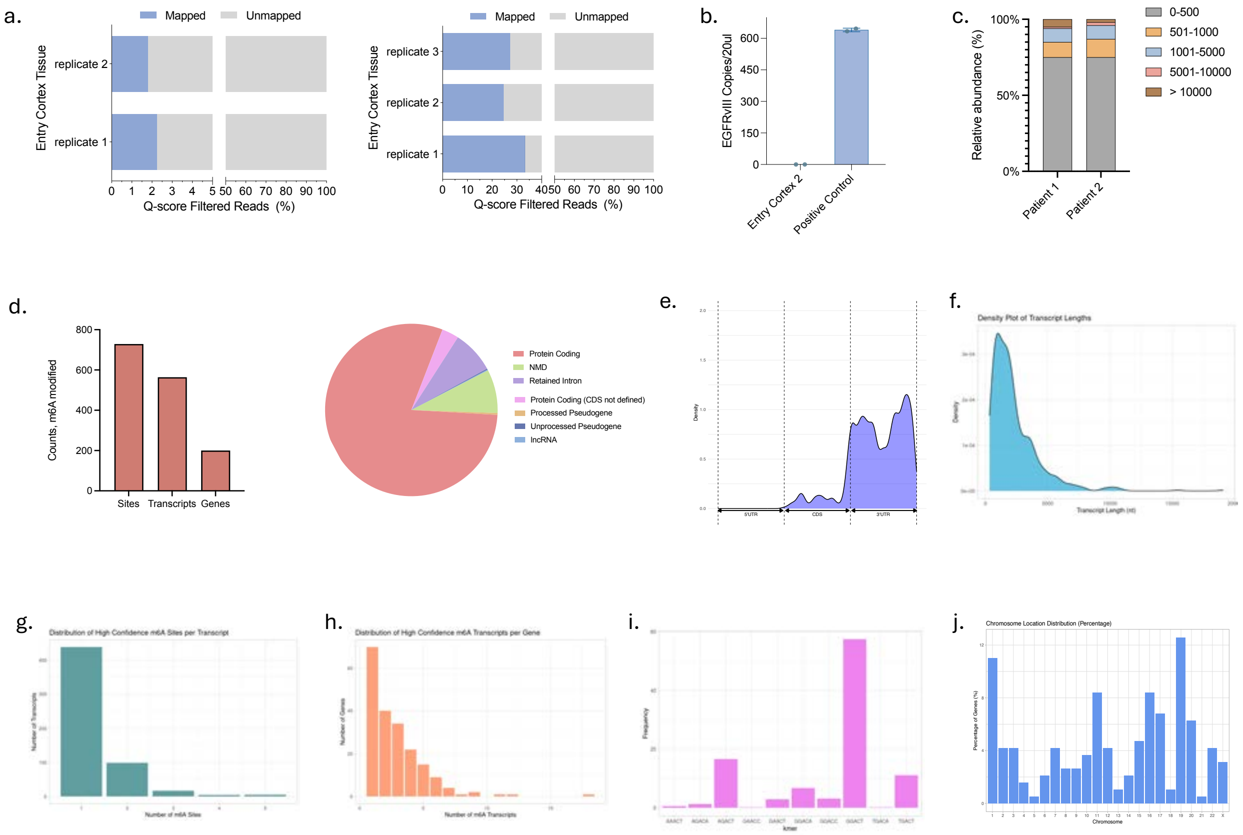

**Supplementary Figure 9. Entry Cortex Sequencing Output and m6A Modification Metrics.** (a) Distribution of mapped and unmapped reads from Entry Cortex Samples after alignment, with Patient 2 (right, 33.3%, 24.6%, 27.2%) exhibiting a higher mapped reads percentage compared to Patient 1 (left, 2.2%, 1.8%). **(b) …** (c) Stacked bar chart showing the relative abundance (%) of mapped read lengths. Lengths of 0-500 bp were most prevalent (Patient 1 = 75%, Patient 2 = 75%). (d, left) Distribution of high confidence (probability modified > 0.9) detected m6a sites (n=729), transcripts (n=554), and genes (n=200) across pooled replicates for both patients. (d, right) Transcript biotype distribution for modified transcripts, with high prevalence of protein coding (80.0%), followed by nonsense-mediated decay (8.2%), and retained intron (8.0%). (f) KDE plots demonstrating the distribution of modified sites along the transcript regions (5'UTR, CDS, 3'UTR), with peaks at the start and end of the 3'UTR. (f) Density plot of modified transcript lengths with peak at around 1000 nt. (g) Distribution of high confidence m6A sites per transcript (1 site = 439, 2 sites = 99, 3 sites = 17). (h) Distribution of high confidence m6a transcripts per gene (1 transcript = 70, 2 transcripts = 40, 3 transcripts = 34). (i) kmer distribution of high confidence m6a sites with GGACT being the most prevalent (n=419 sites), followed by AGACT (n=121) and TGACT (n=81). (j) Chromosomal distribution of modified genes with peaks at chromosome 1 (n = 21, 11.0%) and 19 (n=24, 12.6%).

#### Supplementary Figure 10

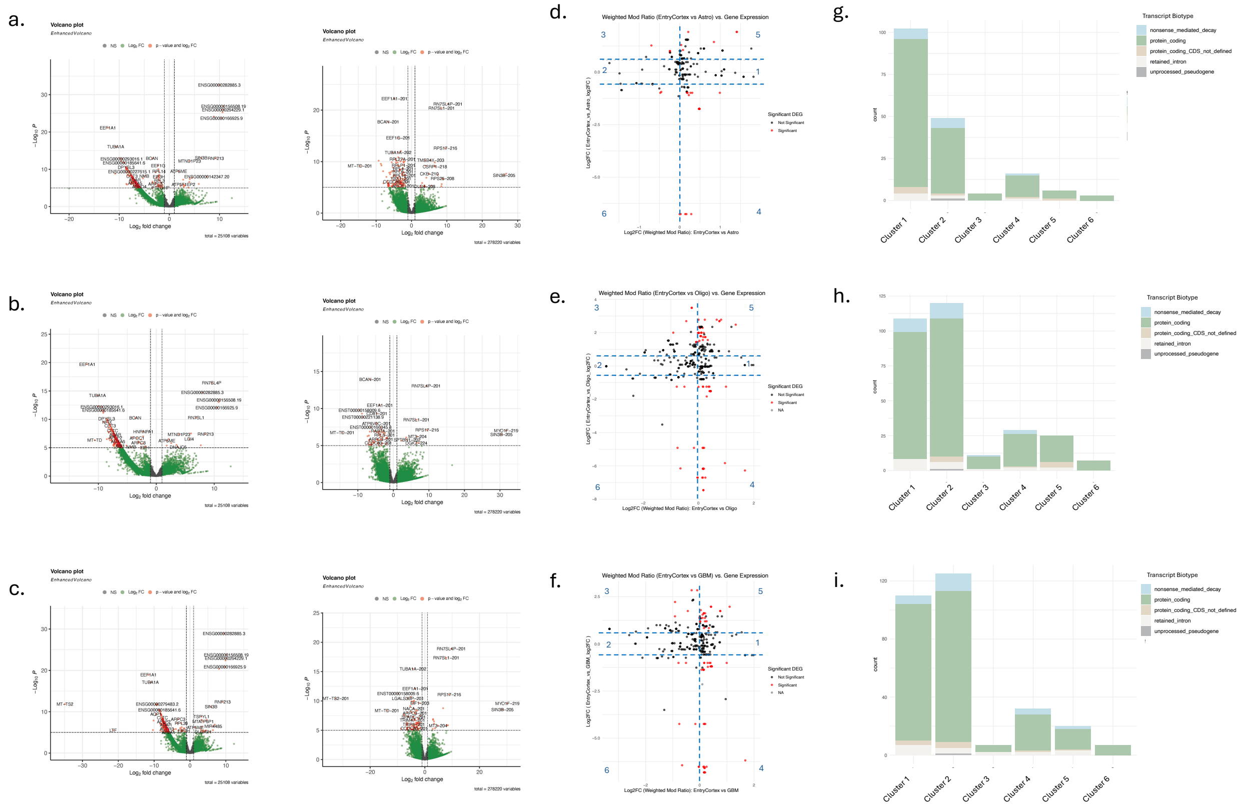

**Supplementary Figure 10. Differential Expression and m6A Methylation between Glioma and Entry Cortex.**
(a-c) Volcano plots depicting differential gene (left) and transcript (right) expression between Glioma and Entry Cortex samples (a, Astrocytoma vs Entry Cortex, b, Oligodendroglioma vs Entry Cortex; c, GBM vs Entry Cortex) Positive log2 fold change indicates upregulation in Entry Cortex and negative log2 fold change indicates upregulation in glioma. Significant changes in gene and transcript expression are highlighted in red. (d-f) Scatter plots illustrating the differential methylation vs. gene expression analysis of common m6A modified genes across glioma subtypes and Entry Cortex. The x-axis represents the log2 fold change (FC) in m6A methylation levels, while the y-axis denotes the log2 FC of differentially expressed genes (DEGs). DEGs with Log2 FC > 0.58 or < -0.58 and p-value < .05 are highlighted in red, while non-significant ones are shown in black. The analysis is performed across three comparisons: Astrocytoma vs. Entry Cortex (d), Oligodendroglioma vs. Entry Cortex (e), and GBM vs. Entry Cortex. Clusters of hyper- and hypo- methylation and up- and down- regulated m6A genes are labeled. (e) RNA biotype distributions are graphed for clusters in the three study comparisons: (g) Astrocytoma vs. GBM, (h) Oligodendroglioma vs. GBM, (i) Astrocytoma vs. Oligodendroglioma.

#### Supplementary Figure 11

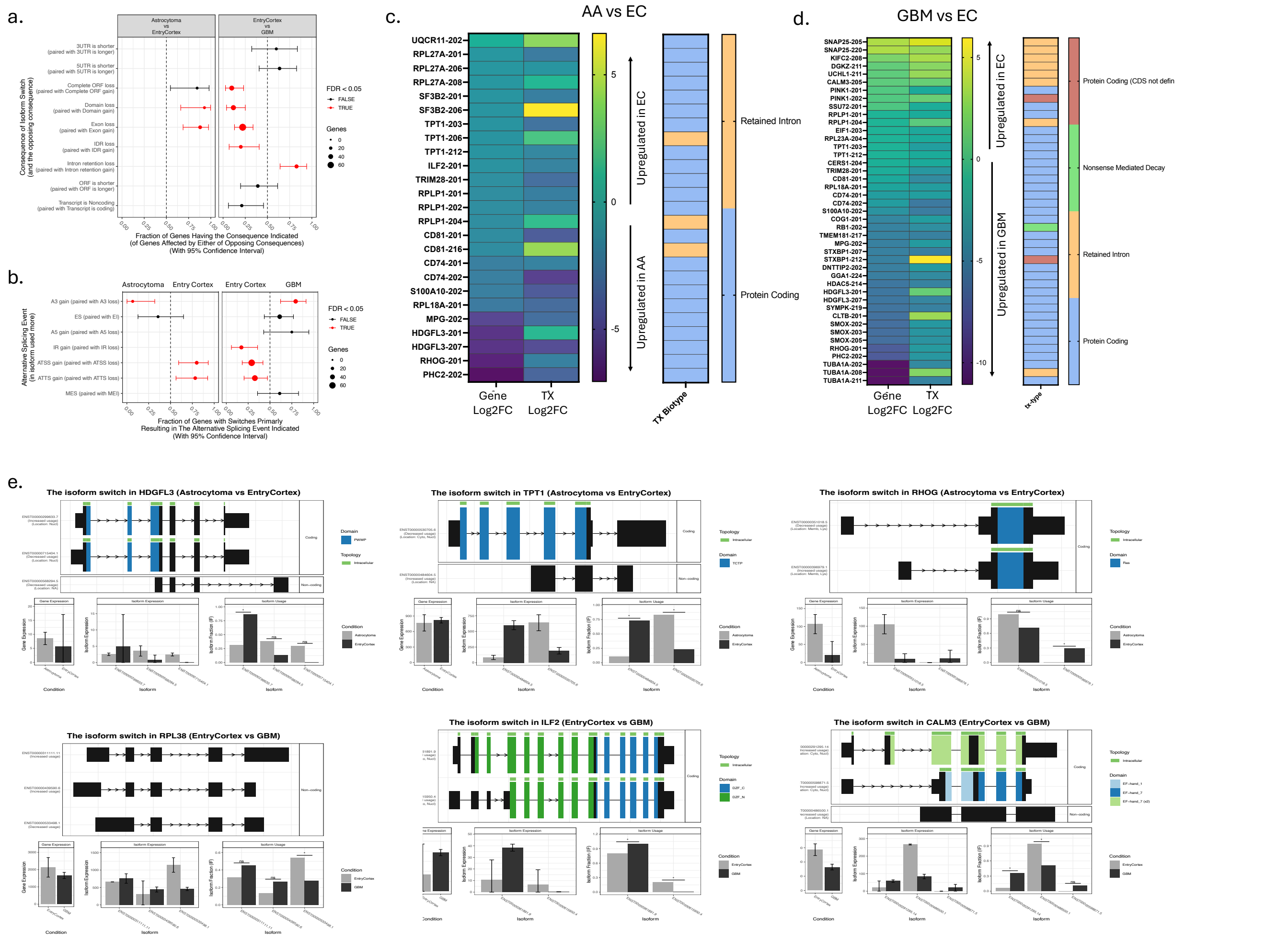

**Supplementary Figure 11: Isoform Usage Analysis between Glioma and Entry Cortex**
(a) Consequence enrichment analysis identifying predicted structural consequences of isoform switches between Astrocytoma vs Entry Cortex (left) and Entry Cortex vs GBM (right). Y-axis: consequence, x-axis: fraction of genes having that consequence (95% confidence interval, CI). Consequences with a fraction of genes greater than 0.5 indicate higher consequence prevalence in Entry Cortex (Astrocytoma vs Entry Cortex) and in GBM (Entry Cortex vs GBM). Significant consequence differences are highlighted in red. Dot size correlates with the number of genes with that consequence. (b) Alternative splicing event enrichment analysis in AA vs GBM. (c-d) Heatmap of the significantly differentially expressed genes (Gene log2FC) and differentially expressed transcripts (TX Log2FC) with significant difference in Isoform usage for Astrocytoma vs Entry Cortex (c) and GBM vs Entry Cortex (d). Transcript biotype annotations are provided. Asterisks indicate common differentially expressed genes between the two comparisons. (e) Switch Plots displaying the isoform structures with annotations, expression levels, and isoform usage.

#### Supplementary Figure 12

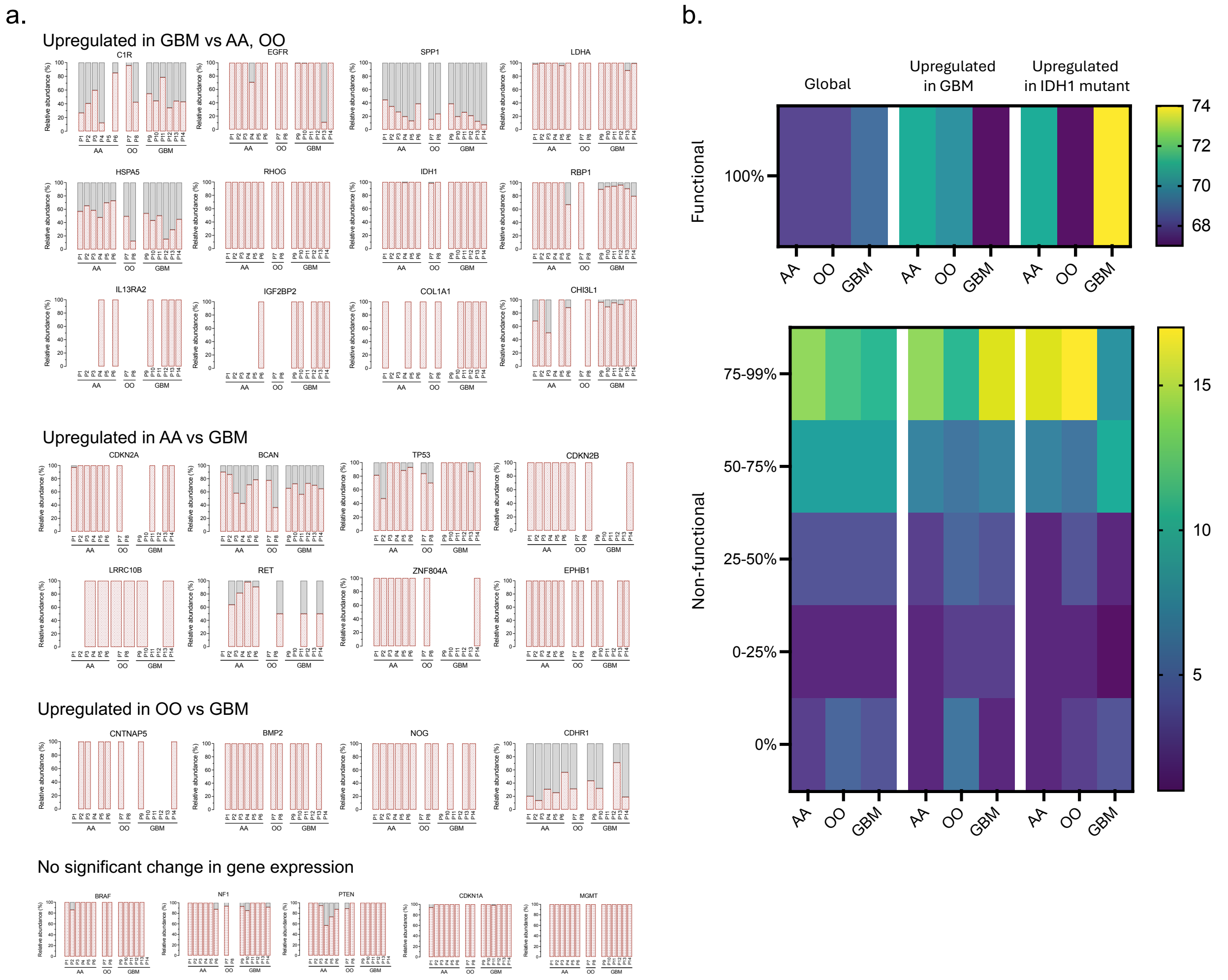

**Supplementary Figure 12. Analysis of functional and non-functional transcript composition among protein-coding genes.**

(a) Relative abundance of functional and non-functional transcripts per gene across patients. Protein-coding genes were stratified based on differential expression: genes upregulated in GBM compared to AA and OO, genes upregulated in AA versus GBM, genes upregulated in OO versus GBM, and genes with no significant differential expression. (b) Heatmap showing the percentage distribution of genes containing 0–100% functional transcripts. The left panel includes all expressed genes, the middle panel highlights genes upregulated in GBM relative to IDH1 mutant gliomas, and the right panel shows genes upregulated in IDH1 mutant gliomas compared to GBM.

**Supplementary Figure 13.**

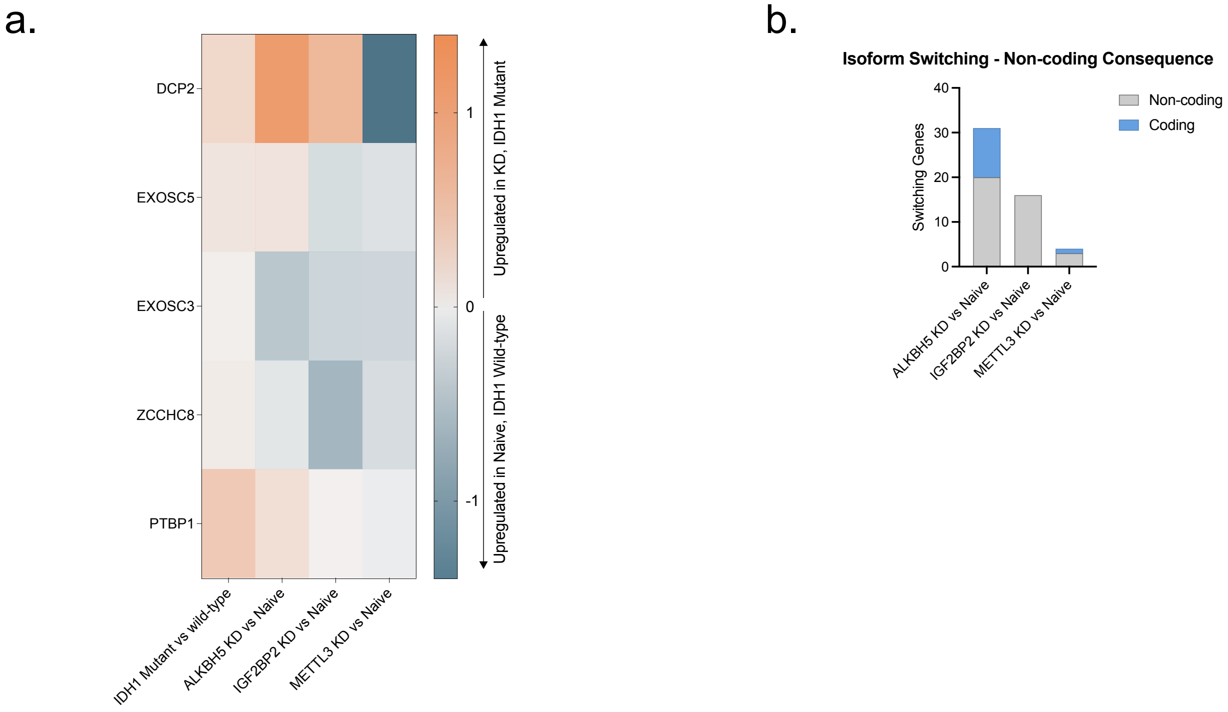

**Supplementary Figure 13: Validation with Knockdown Regulations in Glioma Cell Lines.** (a) Heatmap showing selected splicing and decay factors with differential expression across four comparisons: IDH1 mutant vs IDH1 wild-type glioma tissue, ALKBH5 knockdown vs Naïve, IGF2BP2 knockdown vs Naïve, and METTL3 knockdown vs Naïve. Orange indicates higher expression in IDH1 mutant and knockdown conditions; blue indicates higher expression in Naïve and IDH1 wild-type conditions. (b) Stacked bar chart showing the number of genes with isoform switches producing coding or non-coding consequences in three cell line comparisons: ALKBH5 knockdown vs Naïve, IGF2BP2 knockdown vs Naïve, and METTL3 knockdown vs Naïve.

### Supplementary Table 1

Distribution of RNA biotypes and m6A localization on transcript for hyper- and hypo- methylated transcripts, grouped by the direction of the differential gene expression. Three comparisons are shown: AA vs GBM, OO vs GBM, AA vs OO.

#### GENES UPREGULATED IN AA (VS GBM)

| Genes Upreg | Freq | | | | | |
|---|---|---|---|---|---|---|
| 4 | 0.06153846 | TX Hypermeti | Freq | TX Biotype | TX | Freq |
| | | 19 | 0.86363636 | Protein Codi | 15 | 0.78947368 |
| | | | | Retained Intr | 4 | 0.21052632 |
| | | | | NMD | | |
| | | TX Hypermeti | Freq | TX Biotype | TX | Freq |
| | | 3 | 0.13636364 | Protein Codi | 3 | 1 |
| | | | | Retained Intron | | |
| | | | | NMD | | |

Astro m6A Sites

| | | 3UTR | CDS | 5UTR | Total TX | 19 |
|---|---|---|---|---|---|---|
| Hypermethylated in AA | Protein Codi | 24 | 3 | 0 | Total Sites | 35 |
| | Retained Intr | 0 | 0 | 8 | | |
| | NMD | | | | | |
| | | 3UTR | CDS | 5UTR | Total TX | 3 |
| Hypermethylated in GBM | Protein Codi | 5 | 1 | 0 | Total Sites | 6 |
| | Retained Intron | | | | | |
| | NMD | | | | | |

GBM m6A Sites

| | | 3UTR | CDS | 5UTR | Total TX | 19 |
|---|---|---|---|---|---|---|
| Hypermethylated in AA | Protein Codi | 24 | 3 | 0 | Total Sites | 35 |
| | Retained Intr | 0 | 0 | 8 | | |
| | NMD | | | | | |
| | | 3UTR | CDS | 5UTR | Total TX | 3 |
| Hypermethylated in GBM | Protein Codi | 5 | 1 | 0 | Total Sites | 6 |
| | Retained Intron | | | | | |
| | NMD | | | | | |

#### GENES UPREGULATED IN AA (VS GBM)

| Genes Upreg | Freq | | | | | |
|---|---|---|---|---|---|---|
| 11 | 0.16923077 | TX Hypermeti | Freq | TX Biotype | | Freq |
| | | 21 | 0.6 | Protein Codi | 18 | 0.85714286 |
| | | | | Retained Intron | | |
| | | | | NMD | 3 | 0.14285714 |
| | | TX Hypermeti | Freq | TX Biotype | | Freq |
| | | 14 | 0.4 | Protein Codi | 4 | 0.28571429 |
| | | | | Retained Intr | 8 | 0.57142857 |
| | | | | NMD | 2 | 0.14285714 |

Astro m6A Sites

| | | 3UTR | CDS | 5UTR | NA | Total TX | 21 |
|---|---|---|---|---|---|---|---|
| Hypermethylated in AA | Protein Codi | 11 | 17 | 0 | | Total Sites | 33 |
| | Retained Intron | | | | | | |
| | NMD | 5 | 0 | 0 | | | |
| | | 3UTR | CDS | 5UTR | NA | Total TX | 14 |
| Hypermethylated in GBM | Protein Codi | 4 | 3 | 0 | | Total Sites | 25 |
| | Retained Intr | 0 | 0 | 13 | 1 | | |
| | NMD | 3 | 1 | | | | |

GBM m6A Sites

| | 3UTR | CDS | 5UTR | NA | Total TX | 21 |
|---|---|---|---|---|---|---|
| Protein Codi | 9 | 16 | 0 | | Total Sites | 30 |
| Retained Intron | | | | | | |
| NMD | 5 | 0 | 0 | | | |
| | GBM | | | | | |
| | 3UTR | CDS | 5UTR | NA | Total TX | 14 |
| Protein Codi | 5 | 3 | 0 | | Total Sites | 30 |
| Retained Intr | 0 | 0 | 17 | 1 | | |
| NMD | 3 | 1 | | | | |

#### No Significant Gene Expression Difference

| No Significant Gene Expression Difference | Freq | | | | | |
|---|---|---|---|---|---|---|
| 49 | 0.75384615 | TX Hypermeti | Freq | TX Biotype | | Freq |
| | | 128 | 0.75739645 | Protein Codi | 104 | 0.8125 |
| | | | | Retained Intr | 12 | 0.09375 |
| | | | | NMD | 7 | 0.0546875 |
| | | | | Protein Codi | 4 | 0.03125 |
| | | | | Unprocessed | 1 | 0.0078125 |
| | | TX Hypermeti | Freq | TX Biotype | | Freq |
| | | 41 | 0.24260355 | Protein Codi | 35 | 0.85365854 |
| | | | | Retained Intr | 4 | 0.09756098 |
| | | | | NMD | 1 | 0.02439024 |
| | | | | Mt_RNA | 1 | 0.02439024 |

Astro m6A Sites

| | | 3UTR | CDS | 5UTR | NA | Total TX | 128 |
|---|---|---|---|---|---|---|---|
| Hypermethylated in AA | Protein Codi | 133 | 49 | 0 | | Total Sites | 215 |
| | Retained Intr | 0 | 0 | 9 | 6 | | |
| | NMD | 8 | 4 | | | | |
| | Protein Coding (CDS not defined) | | | 3 | 2 | | |
| | Unprocessed Pseudogene | | | | 1 | | |
| | | 3UTR | CDS | 5UTR | NA | Total TX | 41 |
| Hypermethylated in GBM | Protein Codi | 44 | 5 | 0 | | Total Sites | 57 |
| | Retained Intron | | | 4 | 2 | | |
| | NMD | 1 | | | | | |
| | Mt_RNA | | | | 1 | | |

GBM m6A Sites

| | 3UTR | CDS | 5UTR | NA | Total TX | 128 |
|---|---|---|---|---|---|---|
| Protein Codi | 96 | 32 | 0 | | Total Sites | 156 |
| Retained Intr | 0 | 0 | 9 | 6 | | |
| NMD | 6 | 2 | | | | |
| Protein Coding (CDS not defined) | | | 3 | 1 | | |
| Unprocessed Pseudogene | | | | 1 | | |
| | 3UTR | CDS | 5UTR | NA | Total TX | 41 |
| Protein Codi | 45 | 9 | 0 | | Total Sites | 62 |
| Retained Intron | | | 4 | 2 | | |
| NMD | 1 | | | | | |
| Mt_RNA | | | | 1 | | |

#### GENES UPREGULATED IN OO (VS GBM)

| Genes Upreg | Freq | | | | | |
|---|---|---|---|---|---|---|
| 1 | 0.01538462 | TX Hypermeti | Freq | | | Freq |
| | | 1 | 1 | Protein Codi | 1 | 1 |
| | | | | Retained Intron | | |
| | | | | NMD | | |
| | | TX Hypermeti | Freq | | | Freq |
| | | | 0 | Protein Coding | | 0 |
| | | | | Retained Intron | | |
| | | | | NMD | | |

OO m6A Sites

| | | 3UTR | CDS | 5UTR | Total TX | 1 |
|---|---|---|---|---|---|---|
| Hypermethylated in OO | Protein Coding | | 1 | | Total Sites | 1 |
| | Retained Intron | | | | | |
| | NMD | | | | | |
| | | 3UTR | CDS | 5UTR | Total TX | |
| Hypermethylated in GBM | Protein Coding | | | | Total Sites | |
| | Retained Intron | | | | | |
| | NMD | | | | | |

GBM m6a Sites

| | 3UTR | CDS | 5UTR | Total TX | 1 |
|---|---|---|---|---|---|
| Protein Coding | | 1 | | Total Sites | 1 |
| Retained Intron | | | | | |
| NMD | | | | | |
| | 3UTR | CDS | 5UTR | Total TX | |
| Protein Coding | | | | Total Sites | |
| Retained Intron | | | | | |
| NMD | | | | | |

#### GENES UPREGULATED IN GBM (VS OO)

| Genes Upreg | Freq | | | | | |
|---|---|---|---|---|---|---|
| 11 | 0.16923077 | TX Hypermeti | Freq | | | Freq |
| | | 25 | 0.73529412 | Protein Codi | 15 | 0.6 |
| | | | | Retained Intr | 7 | 0.28 |
| | | | | NMD | 2 | 0.08 |
| | | | | Protein Codi | 1 | 0.04 |
| | | TX Hypermeti | Freq | | | Freq |
| | | 10 | 0.29411765 | Protein Codi | 9 | 0.9 |
| | | | | Retained Intr | 1 | 0.1 |
| | | | | NMD | | |

OO m6A Sites

| | | 3UTR | CDS | 5UTR | NA | Total TX | 25 |
|---|---|---|---|---|---|---|---|
| Hypermethylated in OO | Protein Codi | 13 | 9 | 0 | | Total Sites | 41 |
| | Retained Intron | | | 16 | | | |
| | NMD | 2 | | | | | |
| | Protein Coding (CDS not defined) | | | **1** | | | |
| | | 3UTR | CDS | 5UTR | NA | Total TX | 9 |
| Hypermethylated in GBM | Protein Codi | 9 | | | | Total Sites | 12 |
| | Retained Intron | | | 3 | | | |
| | NMD | | | | | | |

GBM m6A Sites

| | 3UTR | CDS | 5UTR | NA | Total TX | 25 |
|---|---|---|---|---|---|---|
| Protein Codi | 12 | 6 | 0 | | Total Sites | 35 |
| Retained Intron | | | 13 | | | |
| NMD | 2 | | | | | |
| Protein Coding (CDS not defined) | | | 1 | | | |
| | 3UTR | CDS | 5UTR | NA | Total TX | 9 |
| Protein Codi | 10 | | | | Total Sites | 13 |
| Retained Intron | | | 3 | | | |
| NMD | | | | | | |

#### No Significant Gene Expression Difference

| No Significant Gene Expression Difference | Freq | | | | | |
|---|---|---|---|---|---|---|
| 53 | 0.81538462 | TX Hypermeti | Freq | | | Freq |
| | | 140 | 0.73684211 | Protein Codi | 112 | 0.8 |
| | | | | Retained Intr | 15 | 0.10714286 |
| | | | | NMD | 9 | 0.06428571 |
| | | | | Protein Codi | 2 | 0.01428571 |
| | | | | Unprocessed | 1 | 0.00714286 |
| | | | | MT-RNA | 1 | 0.00714286 |
| | | TX Hypermeti | Freq | | | Freq |
| | | 50 | 0.26315789 | Protein Codi | 42 | 0.84 |
| | | | | Retained Intr | 5 | 0.1 |
| | | | | NMD | 2 | 0.04 |
| | | | | Protein Codi | 1 | 0.02 |

OO m6A Sites

| | | 3UTR | CDS | 5UTR | NA | Total TX | 140 |
|---|---|---|---|---|---|---|---|
| Hypermethylated in OO | Protein Codi | 182 | 74 | 1 | | Total Sites | 303 |
| | Retained Intron | | | 18 | 7 | | |
| | NMD | 11 | 4 | 0 | | | |
| | Protein Coding (CDS not defined) | | | 1 | 3 | | |
| | Unprocessed Pseudogene | | | | 1 | | |
| | MT-RNA | | | | 1 | | |
| | | 3UTR | CDS | 5UTR | NA | Total TX | 50 |
| Hypermethylated in GBM | Protein Codi | 54 | 3 | | | Total Sites | 67 |
| | Retained Intron | | | 3 | 3 | | |
| | NMD | 3 | | | | | |
| | Protein Coding (CDS not defined) | | | 1 | | | |

GBM m6a Sites

| | 3UTR | CDS | 5UTR | NA | Total TX | 140 |
|---|---|---|---|---|---|---|
| Protein Codi | 107 | 53 | 0 | | Total Sites | 200 |
| Retained Intron | | | 18 | 6 | | |
| NMD | 9 | 3 | 0 | | | |
| Protein Coding (CDS not defined) | | | 1 | 1 | | |
| Unprocessed Pseudogene | | | | 1 | | |
| MT-RNA | | | | 1 | | |
| | 3UTR | CDS | 5UTR | NA | Total TX | 50 |
| Protein Codi | 55 | 4 | | | Total Sites | 71 |
| Retained Intron | | | 4 | 3 | | |
| NMD | 4 | | | | | |
| Protein Coding (CDS not defined) | | | 1 | | | |

#### GENES UPREGULATED IN AA (VS OO)

| Genes Upreg | Freq | | | | | |
|---|---|---|---|---|---|---|
| 4 | | TX Hypermeti | Freq | | | Freq |
| | | 8 | 0.66666667 | Protein Codi | 5 | |
| | | | | Retained Intr | 2 | |
| | | | | Protein Codi | 1 | |
| | | TX Hypermeti | Freq | | | Freq |
| | | 4 | 0.33333333 | Protein Codi | 3 | |
| | | | | Retained Intr | 1 | |

Astro m6A Sites

| | | 3UTR | CDS | 5UTR | Total TX | 8 |
|---|---|---|---|---|---|---|
| Hypermethylated in AA | Protein Codi | 6 | | | Total Sites | 9 |
| | Retained Intron | | | 2 | | |
| | NMD | | | 1 | | |
| | | 3UTR | CDS | 5UTR | Total TX | 4 |
| Hypermethylated in OO | Protein Codi | 3 | | | Total Sites | 4 |
| | Retained Intron | | | 1 | | |
| | NMD | | | | | |

OO m6A Sites

| | 3UTR | CDS | 5UTR | Total TX | 8 |
|---|---|---|---|---|---|
| Protein Codi | 5 | | | Total Sites | 8 |
| Retained Intron | | | 2 | | |
| NMD | | | 1 | | |
| | 3UTR | CDS | 5UTR | Total TX | 4 |
| Protein Codi | 3 | | | Total Sites | 5 |
| Retained Intron | | | 2 | | |
| NMD | | | | | |

#### GENES UPREGULATED IN OO (VS AA)

| Genes Upreg | Freq | | | | | |
|---|---|---|---|---|---|---|
| 2 | | TX Hypermeti | Freq | | | Freq |
| | | 9 | | Protein Codi | 9 | |
| | | | | Retained Intron | | |
| | | | | NMD | | |
| | | | | Protein Coding (CDS not defined) | | |
| | | TX Hypermeti | Freq | | | Freq |
| | | 2 | | Protein Codi | 2 | |
| | | | | Retained Intron | | |
| | | | | NMD | | |

Astro m6A Sites

| | | 3UTR | CDS | 5UTR | NA | Total TX | 9 |
|---|---|---|---|---|---|---|---|
| Hypermethylated in AA | Protein Codi | 12 | | | | Total Sites | 12 |
| | Retained Intron | | | | | | |
| | NMD | | | | | | |
| | Protein Coding (CDS not defined) | | | | | | |
| | | 3UTR | CDS | 5UTR | NA | Total TX | 2 |
| Hypermethylated in OO | Protein Codi | 7 | | | | Total Sites | 7 |
| | Retained Intron | | | | | | |
| | NMD | | | | | | |

OO m6A Sites

| | 3UTR | CDS | 5UTR | NA | Total TX | 9 |
|---|---|---|---|---|---|---|
| Protein Codi | 11 | | | | Total Sites | 11 |
| Retained Intron | | | | | | |
| NMD | | | | | | |
| Protein Coding (CDS not defined) | | | | | | |
| | 3UTR | CDS | 5UTR | NA | Total TX | 2 |
| Protein Codi | 7 | | | | Total Sites | 7 |
| Retained Intron | | | | | | |
| NMD | | | | | | |

#### No Significant Gene Expression Difference

| No Significant Gene Expression Difference | Freq | | | | | |
|---|---|---|---|---|---|---|
| 59 | 0.90769231 | TX Hypermeti | Freq | | | Freq |
| | | 76 | | Protein Codi | 62 | |
| | | | | Retained Intr | 10 | |
| | | | | NMD | 3 | |
| | | | | Protein Codi | 1 | |
| | | TX Hypermeti | Freq | | | Freq |
| | | 127 | | Protein Codi | 98 | |
| | | | | Retained Intr | 15 | |
| | | | | NMD | 10 | |
| | | | | Mt-RNA | 1 | |
| | | | | Protein Codi | 2 | |
| | | | | Unprocessed | 1 | |

Astro m6A Sites

| | | 3UTR | CDS | 5UTR | NA | Total TX | 76 |
|---|---|---|---|---|---|---|---|
| Hypermethylated in AA | Protein Codi | 81 | 26 | | | Total Sites | 131 |
| | Retained Intron | | | 11 | 4 | | |
| | NMD | 5 | 3 | | | | |
| | Protein Coding (CDS not defined) | | | 1 | | | |
| | Unprocessed Pseudogene | | | | | | |
| | MT-RNA | | | | | | |
| | | 3UTR | CDS | 5UTR | NA | Total TX | 127 |
| Hypermethylated in OO | Protein Codi | 112 | 52 | | | Total Sites | 208 |
| | Retained Intron | | | 20 | 5 | | |
| | NMD | 12 | 2 | | | | |
| | Mt-RNA | | | | 1 | | |
| | Protein Coding (CDS not defined) | | | 1 | 2 | | |
| | Unprocessed Pseudogene | | | | 1 | | |

OO m6A Sites

| | 3UTR | CDS | 5UTR | NA | Total TX | 76 |
|---|---|---|---|---|---|---|
| Protein Codi | 75 | 20 | | | Total Sites | 115 |
| Retained Intron | | | 10 | 4 | | |
| NMD | 4 | 1 | | | | |
| Protein Coding (CDS not defined) | | | 1 | | | |
| Unprocessed Pseudogene | | | | | | |
| MT-RNA | | | | | | |
| | 3UTR | CDS | 5UTR | NA | Total TX | 127 |
| Protein Codi | 157 | 67 | 1 | | Total Sites | 278 |
| Retained Intron | | | 26 | 6 | | |
| NMD | 12 | 3 | | | | |
| Mt-RNA | | | | 1 | | |
| Protein Coding (CDS not defined) | | | 1 | 3 | | |
| Unprocessed Pseudogene | | | | 1 | | |
